# Decolonising Global Health: a scoping review

**DOI:** 10.1101/2025.03.26.25324588

**Authors:** Daniel E Stewart, Tiruneh Amsalu, Ellie Fairfoot, David Keen, Jessica Keenan, Frances Butcher, Kevin Miles, Ahmed Razavi

## Abstract

Though much has been written about the importance of decolonising global health, there is a lack of consensus around how it should be defined, conceptualised and actioned, and who has responsibility to do so. In accordance with PRISMA guidelines, we undertook a scoping review of the decolonising global health literature to explore the meaning of decolonising global health, to identify examples of best practice, and to find out how those writing about the issue see the future of the movement.

We searched databases for peer-reviewed and grey literature with titles and abstracts, and then full texts double-screened by authors to identify papers for inclusion. Our search strategy focussed on opinions and discourse using terms broadly linked to decolonising global health. Papers published in either the peer reviewed and grey literature were eligible for inclusion. Data, including conclusions and recommendations, were extracted and results presented as a narrative synthesis of included papers to provide a contemporary account of the decolonising global health agenda.

Included papers (n=129) were predominantly commentary or opinion pieces (n=95). Authors of the included papers were affiliated with institutions predominantly from high income countries including the USA (n=53) and UK (n=30). Included papers presented a broad range of definitions for decolonising global health, describe the historical, colonial influence on global health, explore power imbalances in current global health structures, and make a number of suggestions as to how to address these imbalances.

Despite the clear imperative in the literature to take action, there is no clear consensus on where to start. Drawing from the findings of our review, we conclude with a set of recommended approaches and next steps for decolonising global health, focussing on epistemic injustice, partnership working, the structure of global health, and individual duty.

## 1. INTRODUCTION AND BACKGROUND

Global health actors and institutions must hold the principles of equity and justice at the centre of what they set out to achieve. However, there is much work to be done. The international response to the COVID-19 pandemic has, to a large extent, reinforced pre-existing structural inequities and injustices.

A key injustice relates to the continuing legacy of our colonial past. The decolonising global health agenda recognises that the way in which global health is taught, practised and implemented often perpetuates historically rooted and exploitative power structures, and seeks to address this.

Though much has been written about the importance of decolonising global health in the peer-reviewed and grey literature, the only other review, to our knowledge, focuses specifically on decolonising global health evaluation [1]. This review responds to a gap in the literature about conceptualising the decolonising health movement by exploring what is written in the peer reviewed and grey literature about decolonising global health. This review assesses the literature through identifying the key themes and to generate a set of actions for global health actors and institutions looking to take a decolonising approach.

Our specific aims were to:

a. Collate and critically appraise publications focussed on decolonising global health;
b. Explore themes within the literature, including the conceptualisation and definitions of decolonising global health, examples of best practice actions to decolonise global health, and the challenges to and opportunities for action on this agenda;
c. Generate a set of proposals with a view to direct future research and action focussed on decolonising global health.

## 2. METHODS

We undertook a scoping review of the literature in accordance with PRISMA guidelines for scoping reviews [2] and guided by the methodological framework proposed by Arksey and O’Malley[3]. Scoping reviews are well suited to providing an overview of evidence and for clarifying concepts and definitions [4]. The protocol for this study, registered prior to its undertaking, can be found here: https://osf.io/ynpkf/

### Eligibility criteria and search strategy

Any study or paper type was included in our review including publications both in the peer reviewed and grey literature. Papers published in the English language only were included with no date restrictions on publication date applied.

Papers which directly addressed or explored decolonising global health, including discussions around how it is defined, conceptualised and put into practice were eligible for inclusion.

Our search strategy (S1 Appendix) was adapted to the literature, which was predominantly focussed on opinions and discourse, rather than, for example, intervention studies. The phrase ‘decolonising global health’ was too narrow as literature on the topic is frequently not labelled in this way. As an alternative, we used a broad set of terms, applying a system filter for letters and editorials as these were most likely to express views and opinions. In addition, a term filter in the form of a search string was also used to narrow the results. All terms were searched in abstracts, keywords, subject headings, titles and text words.

We searched Embase, EBSCO Global Health, Medline and Scopus bibliographic databases. A grey literature search using Google search and Google Scholar was also undertaken to capture opinion pieces that may not be indexed by the bibliographic databases. We also undertook citation searches, using a number of source articles which we identified from the search and through our own reading [5–14]. To ensure the literature we included was as up to date as possible, we undertook searches at two points, the first in September 2022 and the second in September 2024.

Full details of the search strategy, search terms and number of results returned is available in the supplementary material.

### Screening

Search results were de-duplicated using Endnote prior to being screened. The large volume of resulting articles were divided amongst the following authors (DS, TA, EF, DK, JK, KM, and AR). Each title and abstract was independently screened by a minimum of two authors of the above authors using the Rayyan review screening tool [15], and discarded those records which clearly did not meet inclusion criteria. Two authors then independently screened full text articles for inclusion or exclusion. Discrepancies or disagreements were addressed first through discussion and if needed by referral to a third author.

### Data extraction

Data was extracted into a pre-agreed Microsoft Excel form. Data extracted included author, their institutional affiliation, and country in which that institution is based, the date and where published, country or region on which the article was focussed, key conclusions and recommendations for further action. Data were extracted independently for each paper by a minimum of two authors with disparities and disagreements discussed first and referred to a third author if necessary.

### Data synthesis

The approach to data synthesis was guided by the aims of the study and the need to robustly present results that included both a high proportion of commentary and opinion-based articles, but also heterogeneous methods amongst the other articles (see results).

As a consequence of this, no articles were excluded on the basis of quality. This was partly because it was not possible to use a quality assessment tool to assess the heterogeneous articles included, but also because the aim of the scoping review was to collate the full range of what is written about decolonising global health. A risk with applying a quality assessment tool to this literature is that it may inadvertently perpetuate the same biases that the decolonising agenda seeks to address. Khan et al. [6], argue that the decolonisation of academic publishing spaces needs to make room for publications from alternate epistemic standpoints and de-emphasise traditional hierarchies of evidence and practise. This review therefore seeks to meet this aspiration.

Instead, the authors worked together to undertake a narrative synthesis of the findings in all the included articles to provide a contemporary account of the decolonising global health agenda. Narrative synthesis has been shown to be a useful technique in synthesising different types of studies without losing the diversity in study designs and contexts [16]. In this study, the Mindmeister tool was used[17] to discuss, review, map out and agree key themes.

The findings from the included articles are summarised below. The narrative synthesis starts with an analysis of who talks about decolonising global health in the literature and the geographical distribution of authors, describes the colonial legacy of global health and how this affects the current definition of decolonising global health, progresses onto the reasons for decolonising global health, approaches and barriers to this, and finishes with an analysis of the perspectives of low- or middle-income country (LMIC) authors. The discussion then critically analyses this literature to consider gaps, opportunities and future research.

## 3. RESULTS

Electronic database searches identified 1268 records with a further 263 records included through citation searches and further reading (Table 1). Following de-duplication, title and abstract and full-text screening, 129 articles were included in our review (Figure 1: PRISMA flow chart).

**Fig 1:** PRISMA Flow Diagram for the identification of studies and papers for inclusion in the review

**Table 1:** Summary of all papers included in the review.

The majority of the papers included in our review were commentary or opinion pieces (n=95). Other papers were literature or scoping reviews (n=13), qualitative studies (n=15), mixed methods studies (n=4) and case studies (n=2). Authors of the included articles were affiliated with institutions from 56 different countries, with most authors (n=53) affiliated to institutions in the USA, the UK (n=30), and Canada (n=17). Most papers included in our review were published in peer-reviewed journals (n=112), with thirteen papers published in the non-peer reviewed grey literature and four published in books. Articles included in the review were published between 2008 and 2024.

### Who is talking about decolonising global health?

The impact of colonialism on health is far reaching, and as such, has attracted interest and commentary across many sectors. The largest group of authors have written about decolonising health from a health provider perspective. These perspectives include psychology [18], dermatology [19], surgery [20–23], radiology [24], linguistics [25], public health [26–28], epidemiology [29], One Health [30], mental health [31, 32], bioengineering [33], rheumatic heart disease [34], health crises [35], healthcare innovation [36], academic medical centres [37], emergency medicine [38, 39] and global health institutions [40, 41]. Historical perspectives have been drawn upon [42] as have those regarding equity-driven funding [43] and global political economy [44].

Decolonising the research sector has attracted commentary, including research approaches to big data [45, 46], implementation science [47], realist evaluation [48] and perceptions of research fellows [49]. Similarly, research partnerships [7, 26, 50, 51] and publishing [52–54] have drawn critique.

Finally, many papers have focussed on decolonising the education sector, including global health learning [6, 22, 26, 41, 55-63], health care professional education [58, 64, 65] and educational partnerships [66, 67].

### What is the geographical distribution of authors?

Despite the variety of sectors involved in decolonising global health, the location of authors (according to the primary institution that the author is affiliated with at the time of publication) shows a disparity in where the literature is being produced. Far more papers (n=51, 40%) were produced by a single author based in a high-income country (HIC) than by a single author based in a LMIC (as per World Bank classifications) (n=8, 6%). Articles with multiple author locations, produced in collaboration between high-income country (HIC) based authors and LMIC based authors made up 35% (n=45) of the papers. Papers with multiple author locations exclusively from HICs made up 13% (n=16), and papers with multiple author locations exclusively from LMICs made up 6% (n=8). In HIC-LMIC research collaborations, it is also important to consider authorship hierarchies. In these collaborations, 69% (n=47) had a first author based in a HIC; 22% (n=15) had a first author based in an LMIC; and 7% (n=5) had joint first authors based in a HIC and LMIC. Figures 2 and 3 set out a map showing the location of the authors and lead authors of included articles.

**Fig 2:** Location of the authors contributing to papers included in the review* * Definition of author location refers to the primary institution that the author is affiliated with at the time of publication.

**Fig 3:** Location of the first author of papers with multiple authors included in the review** ** This data only includes papers with more than one author. <u>Note</u>: Further details on the numbers of authors from each country are provided at the S2 Appendix.

As also reported by Rees et al. 2024 [68], although there has been an expansion of publications regarding decolonizing global health, the narrative on these topics has primarily been told by authors affiliated with HICs. The reasons for the lack of LMIC authors, and the perspectives from LMIC authors, are explored in the following narrative analysis.

### The legacy of colonial health

The conceptualisation of decolonising global health rests on understanding what colonial health is and the impact of this. The discipline of colonial health developed from European colonialists in the 16th and 17th centuries, who saw health conditions unfamiliar to colonists as a threat to their mission. For instance, by the late 1800s, malaria was considered to be the largest obstacle to colonisation, with metropolitan military and business interests being compromised by the susceptibility of white settlers to malaria, which was by far the largest cause of death for that group [69]. In order for the colonial project to succeed, treatments for diseases impacting colonial administrators as well as the local populations whose labour they exploited were required [70].

In addition to developing strategies to protect the interests of colonisers, the necessity to understand ‘new’ disease aetiologies, patterns, and treatments of the indigenous populations, was usually pitted against the interests of the colonised [71]. Colonial authorities saw themselves as having the right solutions and rarely considered that the people they were trying to ‘help’ could possibly have their own solutions [56]. Most indigenous forms of understanding were devalued and demonised and were soon replaced by ‘technocratic experts’ who showed little concern for the socioeconomic realities on the ground [40, 72, 73]. Epistemic dominance from HICs carved its place. For example, the journal the Lancet was born as a product of colonialism and, at least in part, as an instrument to support and advance British imperial objectives [11].

The legacy of colonial health remains. The creation and dominance of colonial, missionary and tropical medicine followed by international health and now global health, continue to be underpinned by the remnants of colonial heritage and practices [40, 69, 70, 74-78]. These colonial practices have been *“codified into modernity, modern states, economic, social, intellectual and international institutions”*[60] and these institutions continue to reinforce racism and bias [26]. For instance, population refusal to seek care as a result of colonial violence, continues to hamper modern day medical emergencies [79], whilst educational curricula carry these epistemological biases [60], including the exclusion of a ‘Southern’ perspective in intervention design and evaluation research approaches [80]. Tuhebwe and colleagues (2023) argue that colonial power dynamics can be seen throughout the project cycle of many global health programmes.[81]

By ignoring history, and continuing to accept power imbalances, patterns of oppression and exploitation reproduce and support the current system of global health [31]. This includes a distinct power imbalance where ‘resource limitations’ in low-income settings have always been externally imposed [44].

### Defining the decolonising global health movement

The ‘decolonising global health’ movement thus developed from the historical and current legacy of colonial health. Decolonisation has been defined as the elimination of the colonial experience and its legacy [56] to allow for the independence and full agency of all involved organisations, communities, and persons [82]. When applied to global health, the decolonising movement seeks, amongst other things, to acknowledge global health’s roots in colonialism [11]; highlight and challenge the power asymmetries in the global health architecture [83]; remove all forms of supremacy within spaces of global health practice [74]; identify ways in which global health teaching and research can overcome its colonial past [7, 40]; and, advocate for critical global health education grounded in anti-colonial perspectives [83, 84].

Clear distinctions, but links, are also drawn between other similar movements, frameworks and concepts, including the Black Lives Matter movement [83][61], equity, diversity and inclusion (EDI) and anti-racist reform [40, 85]. As such, decolonising global health can also be understood as an approach to social justice also intersects with other harmful ‘-isms’, that pose the largest threat to health equity (e.g. racism, sexism, capitalism) [5]. Decolonising includes anti-racism, not just equality, diversity and inclusion initiatives, but needs to look at the root causes of structural and individual racism to address this issue [86]. The Editors of the Lancet Global Health noted that racist stereotypes have re-emerged towards Africans as a result of COVID-19 [87] demonstrating the intersectional nature of decolonising global health. Nassiri-Ansari and Rhule (2024) emphasise the intersection between race and gender and state that efforts to decolonise global health must focus on both race and gender equality [88].

However, the terminology of decolonising global health is far from universally accepted, lacks clarity [89], is poorly understood [63] and viewed as unconventional [90]. Krugman challenges the *“buzzwordification”* of decolonising global health, emphasizing that decolonisation is a word with underdetermined and contested meanings, associations and representations [91].

The complexity of what it means to decolonise global health should also be acknowledged. Contractor and Dasgupta use the example of Indian’s caste system to draw attention to the local complexities of decolonisation, warning that the Global South is made up of diverse societies and a one-size-fits all approach is not appropriate. They draw attention to the historic cultural imbalances of power which were used in India by the colonisers and which will remain if decolonisation occurs without representation from suppressed minorities [92].

Beyond the discussion over what the terminology of decolonising global health means, there is also a debate over whether the terminology should be used and what alternatives would be preferable. Some authors suggest decolonising global health is defined predominantly by those from HICs [49, 83], predominantly within universities in HICs and therefore are not defined by those at the receiving end of the interventions, thus perpetuating existing power and knowledge structures [7, 40]. For example, Engebretsen critiques the decolonising global health movement, arguing that the rhetoric of decolonising global health has done nothing to address the root causes of the disastrous health situation in Gaza and the West Bank [93].

Hellowell [94] queries whether a decolonising global health framework is the best solution to issues within global health. A problem with this terminology is that it is associated with binary arguments which place people into groups of oppressed and oppressors depending on their background or place of origin. He suggests that solutions derived through this lens have the potential to harm the aims of the global health agenda by: ‘*(i) undermining confidence in scientific knowledge; ii) accentuating inter-group and inter-national antagonisms; and (iii) by discounting the degree of progress already achieved that may curtail opportunities for redistributive change in the future*’.

Binagwaho et al provide an alternative to decolonising global health, arguing that the *“elimination of a white supremacy mindset”* is a better positioned term, as it recognises the crux of colonisation as assumed racial superiority, and how the legacy of such sustains the privilege white people enjoy at the expense of non-white people [56] regardless of colonial roots [95]. Whilst this review uses the decolonising global health terminology throughout, as this was reflected in the search terms and majority of the literature, further exploration of what other terminology is used is included in the discussion.

### Why decolonise global health?

At the heart of why authors advocate that global health must be decolonised is the broad and consistent understanding that while colonisation is largely a remnant of the past, the heritage and legacies of colonialism, such as political and economic structures, healthcare systems, power dynamics, behaviours and partnership inequities continue to permeate across, and be entrenched within, global health [7, 10, 11, 41, 54, 58, 66, 75, 77, 84, 87, 96-99].

The colonial legacy has contributed to the gap in health outcomes [60] and life expectancy [100] between high and low income countries. One enduring problem is that the default patient in the healthcare field is often considered to be a white male, therefore decolonising symptoms, signs and investigations is an important part of why decolonising global health is needed. For instance, white European biochemical and visual norms used universally can increase the risk of misdiagnosis across populations [96].

As such, the argument for decolonising global health comes from the imperative to hold actors, funders and enablers in global health programmes accountable for their (in)action [43], and address ongoing political manipulation, hypocrisy and distrust [101] in order to redefine roles in international partnership [59], and redress injustices and improve health equity [61, 102].

The answer to the question of ‘why decolonise global health now?’ has been shaped by the COVID-19 pandemic in the literature. A number of authors emphasises the important role the COVID-19 pandemic had in highlighting inequities in global health outcomes, partnerships and the delivery of global health programmes, and therefore underlining the need to decolonise global health [71, 84, 103, 104].

### Approaches to decolonising global health

Frameworks, as a means of understanding the need to decolonise global health and as an approach to put a decolonising approach into action were identified in a number of the papers in our search. Amongst the papers included in the more recent of our two searches (September 2024), we noted a greater emphasis on the application and action in the frameworks and approaches identified than amongst the papers identified earlier [24, 38, 39, 105-107].

The frameworks and approaches set out in the papers we included share overlapping themes and approaches. These include utilising an epistemic injustice framework, a focus on ‘true’ partnership working, rethinking the structure of global health, considering individual duties, alongside actions that cut across all these approaches.

#### i) Epistemic injustice

The epistemic injustice framework approach focuses on countering testimonial injustice, specifically where local expertise is excluded from research and local knowledge production is deemed as illegitimate or lesser. It also focuses on countering interpretive injustice by using local interpretive tools and ensuring research aims are not solely aligned to the dominant western audience [45, 61].

The importance of local knowledge and context is consistently highlighted [84]. Avoiding judgement based on foreign cultural norms and using indigenous driven leadership [29, 39, 69, 75, 108, 109] are key to this. Local expert inclusion should be appreciated [24, 38, 50, 51, 84] not just be seen as a tick box [20] and there should be reciprocal knowledge flow [5].

This can be done through mentoring, investing in researchers and agreeing priorities locally [19]. Building up local expertise and supporting existing programmes in LMICs rather than designing programmes around the needs of western students is a way to shift the locus of control [22, 109, 110]. Likewise, medical journals should diversify their boards [19], as the gatekeepers of global health knowledge. Local indigenous knowledge can and should be re-legitimised [30, 73, 108], through reflection on the terminology used when discussing global health challenges [9, 111] and making room for alternate epistemic standpoints [6, 112]. There should also be equity in data collection, analysis, usage and storing [29, 45] to ensure all partners have equitable access, as well as the inclusion of representative participants and researchers in clinical trials, which otherwise leads to a dependence upon clinical guidelines from HIC settings [51].

#### ii) Partnership working

Partnership working is related to epistemic injustice approaches and forms a fundamental part of how the literature suggests approaching decolonising global health, particularly in academic partnerships. Partners should have shared decision making with strategic priorities and implementation driven by those trusted by affected groups [86, 113].

Fundamentally, the locus of control should sit with local institutions rather than ‘expert’ foreign partners [38, 114], abiding by the principle of *“no research about us, without us”* [12, 115]. Through partnership working, communities and global health practitioners based in LMICs should be empowered to shape global health interventions [22, 89, 109, 116]. This is a contrast to global health historically where global health expertise has been concentrated in legacy powers [87] and concerns about the current structure of global health reflecting ‘feudal power’ [77]. This is because funding is suggested to favour HICs with no direct funding to LMICs, instead using ‘feudal intermediaries’ thereby retaining power within the hierarchy of the feudal structure. Dako et al [24]and Kumar et al [105] call for research funding to be more equitably distributed, for greater funder accountability and an end to the donor-driven model. This would entail that funding always includes institutions rather than be structured around local partners being used as sub-grantees, and treatment of researchers should be the same regardless of their origin [95]. Nassiri-Ansari et al [88] concur that greater South-South cooperation in the form of funding and new funding models that favour of multilateralism over bilateral arrangements, operate without stipulations, and respond to locally identified needs would *“shift power from donors to the doers”*.

#### iii) Structure of global health

Many approaches to decolonising global health also focus on rethinking the structure of global health. This can include rethinking and restructuring governance relationships that shape decisions [12, 46, 69, 104, 116] rigorous analysis of power asymmetries [117] and not arbitrary movements [11], and rebuilding research infrastructure from the ground up [10]. Kwete et al. [71] argue that addressing structural issues in global health requires decolonising the political economy first.

This raises the question of systemic issues with how global health interacts with the aid industry. To decolonise global health it is suggested that this needs to start with debt cancellation and non-earmarked budget support [44], fair allocation of resources [66], broadening methods of education and research, changing teaching for western students [22, 84, 110] changing the location of where global health education happens [56], reducing the reliance of HICs health service and research institutions on talent from LMICs [51], stopping the *“brain drain”* of health workers from LMICs [51], and reimagining global health as social medicine [118]. There are also suggestions on how to tackle decolonising global health in academia through diversity in authorship [97, 119], ensuring papers have multilingual abstracts [9] and including study of the place itself as part of the work [79].

#### iv) Individual duty

Though much of the focus on approaches to decolonising global health is systemic and structural, some literature does highlight an individual’s power to change things. Practitioners are asked to reflect and question institutional and structural practices and not to act beyond their expertise [110]. Those engaging in decolonising global health initiatives should be rewarded for their effort and risk in speaking out [86] and not accepting the status quo [110]. This can contribute to building a culture of reflexivity by sparking dialogue in institutions, with the aim of resulting in collective action [120].

It is clear that with such varying systemic, structural and individual issues, decolonising global health will not happen quickly or smoothly. Thus far there has been a highlighting of the need for change and this is a step in the right direction. *“When we make visible this implicit ideological function that the global health field performs, we can see new directions, new ideas and new allies for collective action that are otherwise kept unimaginable”* [121]. This is aspirational, but also requires a pathway to success with action-guiding steps. These could include a commitment to a clear list of reforms to address decolonising global health with metrics to track the progress of these reforms [6, 98] to ensure any action is accountable.

### Barriers to Decolonising Global Health

Implementation of decolonising global health faces multifaceted barriers rooted in both historical legacies and contemporary dynamics.

One critical hurdle is the neglect, at the individual level, to *“emancipate and decolonise our own minds (from the colonial conditionings of our education)”*, a fundamental step often overlooked in the discourse. According to Abimbola et al [55], colonialism sought to infiltrate and manipulate fundamental human values, with the colonial classroom serving as a means of psychological domination in Africa and other regions. They suggest that the lingering effects of colonial education policies have instilled a sense of inferiority in many individuals, highlighting the urgent need to dismantle this mindset and reclaim autonomy over our perceptions and judgments. Failure to address this aspect impedes progress by perpetuating entrenched ideologies and power imbalances.

Araújo et al [50], Farag [122] and Gedela et al [123] believe the undervaluing of science produced in LMICs, particularly by global health leaders, is both an area that the epistemic injustice approach to decolonising global health seeks to address, and an ongoing barrier to decolonising global health. The persistence of such attitudes risks reinforcing disparities and hindering the integration of context-specific solutions into global health initiatives.

Moreover, a dominant perspective in the literature suggests that decolonisation efforts often fall short in addressing what is perceived as the root cause of coloniality: white supremacy. According to Binagwaho et al [56], the global influence of white supremacy, rooted in geographic distinctions and skin colour disparities, pervades various societal domains worldwide. Addressing its presence in global health education is crucial, as its elimination could profoundly benefit the health and welfare of marginalized populations globally. Binagwaho et al [56] and Finkel et al [97] argue that the existing global health discourse inadequately confronts the ongoing manifestation of white supremacy, particularly evident in leadership inequities where white men from HICs dominate key positions and decision-making power is concentrated in HICs. This lack of diversity in senior roles is exacerbated by confusion about the meaning of decolonisation as opposed to inclusion and diversity [86].

Yerramilli [44] also argues that the success of the decolonising global health movement may be dependent on much wider global factors related to decolonising the world’s political economy. This entails dismantling the deep-seated socio-economic inequities exacerbated by historical colonisation. Yerramilli argues that decolonising global health requires us to challenge our concepts of ‘aid’, as this implies voluntary relief, and ‘sustainability’ as this ignores the ongoing political and economic oppression of developing countries. Reimagining the global political economy alongside financial compensation for countries’ historical and ongoing outflows is needed to truly decolonise global health [44] and the overarching influence of the world’s political economy all complicate the path to decolonising global health [10, 43]. Finally, Hellowell 2022 argue that while the decolonisation agenda has the potential to stimulate a much needed redistribution of decision-making power in global health, it also has the potential to undermine confidence in scientific knowledge, accentuating intergroup and international antagonisms and curtailing the opportunities for redistribution in the future [94].

### Perspectives from low and middle income economies

As the importance – and lack – of LMIC voices emerged as a theme in the narrative analysis, we decided to highlight the perspectives of LMIC authors in this scoping review. However, it must be noted that LMIC originating papers generally propose similar concepts and ideas when compared across the spectrum of papers. There are several reasons for this. Mogaka et al [10] suggest researchers from LMICs are educated and working within or toward HIC global health structures and standards. Also, the process by which we identified authors from LMICs, by only referring to papers where authors worked for organisations based in LMICs, may miss or exclude those authors from LMICs who may now reside in HICs [68, 106]. It may be that the search terms used have identified publications by authors who share similar perspectives. And finally, these papers have not been reviewed in isolation and are also embedded within the above narrative.

A particularly strong concern from LMIC authors discussed the disparity of funding and expertise which is perpetuated by the current structures and processes operating across global health [10, 50, 56, 115, 120]. There is a clear demand for parity in partnerships [10, 31, 56] with local expertise and leadership recognised and utilised [31, 99, 115, 120], in a real rather than tokenistic manner [76, 106, 124]. Three papers describe the importance of human factors in decolonisation [71, 120, 125], highlighting that change will not happen without the self-reflection of global health practitioners. Sharma and Sam-Agudu [126] emphasise the particular need for practitioners from the “Global South” to take action to decolonise global health, as the biggest stakeholders and those who stand to benefit the most from decolonising global health. Ssyennyonjo et al. present the decolonising agenda as an opportunity that could be leveraged in order to achieve strategic aspirations defined by Africans themselves [127], through Agenda 2063, a strategic document created by the African Union.[128]

## 4. DISCUSSION

In this paper we have reviewed existing literature around the topic of decolonising global health and reported the range of different discussions and views. We have found that the decolonising global health movement broadly attempts to define what decolonising global health is, explore power imbalances in current global health structures, and how to address these imbalances. Our search showed that the majority of the literature about decolonising global health is in the form of commentary or opinion pieces, with authors based in HICs dominating, although it is difficult to accurately assess every author’s background.

Much of the literature describes the historical origins of global health through colonial medicine and attempts to define decolonising global health in order to frame the discussion on decolonising global health. This then helps define why the decolonising global health movement is needed as a means to address epistemic injustice and reduce health inequalities. Our review suggests that there is no universally accepted definition of decolonising global health, that there is uncertainty over how the term originated and is used to further the interests of HICs, and that there is a question as to whether it is the most appropriate response to the inequities driven by colonialism within global health.

The literature included in our review was generally consistent on approaches to decolonise global health with suggestions including: empowering local communities, increasing opportunities for those in LMICs, addressing epistemic imbalances, moving more decision-making power into LMICs, changing the mindset of the heterosexual, white, European, male being the default reference point for medical practice, and restructuring global health so that leadership and structures are more representative of the global community.

Despite the clear imperative in the literature to take action, there is no clear consensus on where to start and how, even if many of the actions suggested are shared across the literature. As we note, papers identified in the most recent of the two searches of the literature which we undertook more frequently presented actionable next steps. However, where steps are set out (using data and metrics [6], subsidiarity and shared decision making [129], overhauling the global health industry [130]) it is often not clear how these should be coordinated and by whom. This is potentially an outcome of the nascent nature of discussions on how to decolonise global health and further work on this may be forthcoming.

There are clear barriers to decolonising global health that are described within the literature. There is a recognition that much of the imbalance stems from the donor-recipient relationship as a consequence of how aid works in our current global geopolitical system. Indeed, some commentators highlighted that global health in its current format is part of the problem, entrenching imbalances and power dynamics through the use of aid as soft power. Some authors suggested that the structural changes needed are wider than the global health system and extend to looking at the global neo-liberal capitalist system as the root of these inequalities. Despite the prevailing call for structural changes, there appears to have been limited efforts to address these in major global health institutions with only a few exploring how the changes in practice suggested in the literature may be implemented. None of the papers identified in our review reported the results of changes or interventions to implement any such changes in practice. This may be due to a tension between the structural changes called for in global health relying on change to be driven by those leading those very global health institutions that are acknowledged as part of the problem. Indeed, the question remains, who is best placed to drive forward the structural changes in order to make global health more equitable.

In terms of specific actions or principles to decolonise global health, based on the papers appraised in this review, we propose the following as a starting point to further the discussion. These are orientated by the same conceptual classification we saw in the literature on decolonising global health.

### Recommended Approaches

#### Epistemic injustice

1. While it is useful to have a shared terminology and understanding in order to promote good practice, this does not have to be framed as ‘decolonising global health’. Alternative terms from LMIC global health actors should be considered.
2. Academic tools, such as quality assurance frameworks, should be re-engineered to ensure that they reflect and convey a variety of epistemic viewpoints.
3. Global health research, funding, educational and leadership opportunities should be prioritised for global health actors from LMICs to address the importance of local context and knowledge in decision-making.
4. Global health actors, institutions and funders should acknowledge that expertise and knowledge exists and must be considered in the widest range of forms that may include the lived experience, local, cultural, political and traditional knowledge of individuals and communities.

#### Partnership working

1. There is a duty for global health actors to implement and evaluate shared decision making frameworks and processes in global health collaborations.
2. Global health actors should adopt a minimum standard to work towards equitable partnerships, ensuring material and human resources are equitably available to all those involved.
3. When brokering and evaluating global partnerships, global health actors should include assessment of the equity of the partnership.

#### Structure of global health

1. There is a need to generate interventions and metrics to support the decolonisation of global health. These should answer questions such as *‘How do we know how institutions are currently performing?’* or ‘*How do we know that an intervention (e.g. decolonising a learning curriculum) generates the intended outcome and does not accentuate harm.’* There is a need for methodological inquiry and deliberation to think about what metrics would be valid and how progress should be measured.
2. Financial aid flows from HICs to LMICs should be targeted towards LMIC researchers or LMIC-led collaborations.
3. There should be increased diversity and active representation from across LMICs in global health leadership positions.
4. Where organisations commit to decolonising global health, system-wide, organisational and individual accountability mechanisms should be embedded to ensure recommendations are addressed and actions are evaluated.

#### Individual duty

1. Reflective practice in global health should be matched with peer dialogue and insight driven action to better support conditions within organisations to support efforts to decolonise.
2. Global Health actors or those working in or researching global health topics should advocate for decolonising global health to make visible what is at stake.

These recommendations serve, not as an end point, but as a pragmatic starting point for furthering the discussion on how decolonising global health could be driven forward with a view to achieving more equitable outcomes and impact across the sphere of global health. We therefore welcome discussion and critique of these, with the hope that compiling this disparate literature aids in revealing a clearer path forward for this critically important movement.

### Limitations

There are some clear limitations associated with this review. The inclusion criteria focused on papers discussing ‘decolonising global health’ as the primary framework identifier for papers. This may have biased the papers we identified and the views that might be expressed in those papers, as other terms used (possibly in isolation) for the same concept may not have been captured in our search strategy. As we have seen, criticisms of the decolonising global health movement include the idea that this is a conversation that is mostly being driven by HICs and may now be being used as a buzzword to demonstrate equity and justify institutional existence in the global health space to funders and political leaders. It may also be that similar conversations are occurring in the literature without using the specific term ‘decolonising global health’ that we have not therefore captured in this review.

Indeed, the Africa Centres for Disease Control and Prevention, capture a similar concept of centring local communities, ensuring interventions are local priority led and reducing the imbalance of the donor-donee relationship [131]. The term they use for this is ‘the new public health order for Africa’ and the term ‘decolonising global health’ is neither used nor preferred. This is why we, in our recommended approaches, suggest that an LMIC-led consensus on terminology is important, but this terminology does not need to be decolonising global health.

The concept of a ‘new public health order for Africa’ may hint at one of the ways forward for the decolonising global health movement. Movements to strengthen regional health organisations such as Africa CDC and the ASEAN Centre for Public Health Emergencies and Emerging Diseases [132] are key. These organisations emphasise the need for local leadership of priorities, and locally led initiatives to ensure equitable access to interventions such as vaccines. This may be one of the tangible ways that our recommended approach to increasing LMIC leadership and power imbalances within global health may be mitigated.

It is also interesting to note the timing of the proliferation of literature on decolonising global health. Two significant events could be associated with this. Firstly, the Black Lives Matter movement in response to a number of highly publicised cases of US police officers shooting unarmed black people led to an increase in global awareness of social justice with people working in diverse occupations, ranging from health to education [133] to engineering [134] sparking decolonising movements for those sectors. Secondly, the COVID-19 pandemic starkly demonstrated the difference in access to interventions and resources available to different countries [103, 104], especially in terms of personal protective equipment, medical treatment and vaccinations [135]. Calls to decolonise global health often highlighted this inequity as a demonstration of why the decolonising global health movement is needed.

## Supporting information

Fig 1 - PRISMA Flow Diagram

Fig 2 - Author Locations

Fig 3 - Lead Author Locations

Table 1 - Summary of Included Papers

S1 Appendix - Search Strategy

S2 Appendix - Author Locations Tables

## Data Availability

The authors confirm that the data supporting the findings of this study are available within the article and its supplementary materials. Please contact the authors for any questions regarding the data.

## Acknowledgements

The authors would like to thank Anh Tran, Senior Knowledge and Evidence Manager and colleagues at the UK Health Security Agency Knowledge and Library Services for their support and guidance in developing the search strategy for this review.

---

**S1 Appendix: Decolonising global health: A scoping review search strategy**

**S2 Appendix: Country of origin for authors of included papers**

