## Supplementary material for "Decolonising Global Health: a scoping review": Fig 1 - PRISMA Flow Diagram

Records identified through database searches:

- Embase (n = 491)
- Global Health (n = 212)
- Google Scholar (n = 49)
- Google Search (n = 19)
- Medline (n = 670)
- Scopus (n = 948)

Records identified from citation searches:

- citationchaser* (n = 258)

Records identified from wider reading (n=5)

Duplicate records removed before screening:

- Embase (n = 314)
- Global Health (n = 122)
- Google Scholar (n = 3)
- Medline (n = 58)
- Scopus (n = 576)
- citationchaser* (n = 48)

Titles and abstracts screened

(n = 1531)

Records excluded

(n = 1283)

Records sought for full text retrieval

(n = 248)

Records not retrieved

(n = 0)

Full text articles assessed for eligibility

(n = 248)

Full text articles excluded:

- Paper not directly focussed on decolonising global health (n= 88)
- Duplicate (n= 27)
- Paper ineligible – book review (n= 2)
- Paper not in English language (n= 2)

Records included in review

(n = 129)

**Identification**

**Screening**

**Included**

**Identification of studies via databases and registers**

* Haddaway NR, Grainger MJ, Gray CT. Citationchaser: an R package and shiny APP for forward and backward citations chasing in academic searching. Zenodo. 2021.
