## Supplementary figures and images for "Decolonising Global Health: a scoping review"

### Fig 2 - Author Locations

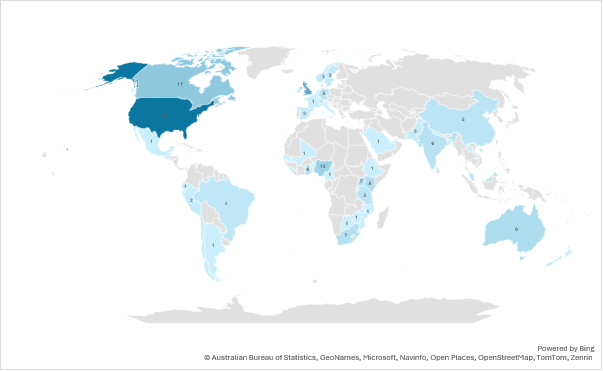

### Fig 3 - Lead Author Locations

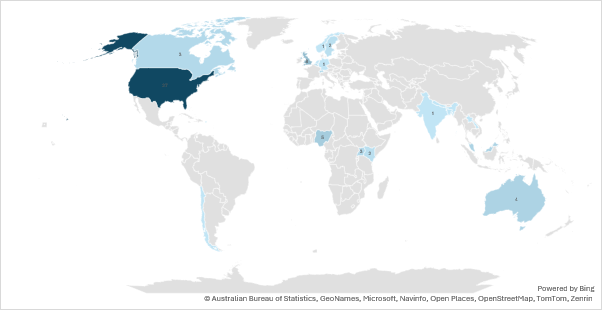
