## Supplementary material for "Decolonising Global Health: a scoping review": Table 1 - Summary of Included Papers

| **Title** | **Year** | **Authors** | **Country** | **Publication Name** | **Type of publication** | **Type of article** | **Key themes** |
| --- | --- | --- | --- | --- | --- | --- | --- |
| 4 Ways to Decolonize Global Health | 2022 | Joanne Silberner | USA | Hopkins Bloomberg Health Magazine | Grey literature | Commentary / Opinion piece | Colonial origins of global health, medical education, reflective practice, individual duty |
| A proposed guide to reducing bias and improving assessments of decolonization in global health research | 2024 | Christine Ngaruiya, Muzzammil Imran Muhammad, Nadia A. Sam-Agudu | USA, Nigeria, Ghana | Frontiers in Education | Journal | Commentary / opinion piece | Moving beyond rhetoric to decolonise global health research, structural change, domains for decolonising global health, power dynamics in global health research, decolonising as an iterative process. |
| A toolkit for decolonizing global emergency medicine education | 2023 | Adeline Dozois, Catalina González Marqués, Kaushila Thilakasiri, Adebisi Anthonia Adeyeye, Joseph Leanza, Megan Rybarczyk, Timothy Depp, Travis Wieland, Naz Karim, Monalisa Muchatuta, Fahad Ali, Ahmed Amer, Stephanie Chow Garbern, Shama Patel | USA, Sri Lanka, Nigeria, | Frontiers in Education | Journal | Mixed methods study | Decolonising global emergency medicine, barriers to global emergency medicine education, cultural humility. |
| Accountability framework to decolonise global health | 2021 | Bolajoko O. Olusanya | Nigeria, South Africa | The Lancet | Journal | Commentary / Opinion piece | Need for systems and frameworks to hold global health institutions accountable |
| Actions for decolonizing global health | 2022 | Garry Aslanyan, Catherine Kyobutungi, Agnes Binagwaho | Canada, Uganda, Rwanda | TDR Global Matters Podcast (transcript) | Grey literature | Commentary / Opinion piece | White supremacy, models of finding for global health, systems change, global health academia |
| Addressing power asymmetries in global health: Imperatives in the wake of the COVID-19 pandemic | 2021 | Seye Abimbola, Sumegha Asthana, Cristian Montenegro, Renzo R. Guinto, Desmond Tanko Jumbam, Lance Louskieter, Kenneth Munge Kabubei, Shehnaz Munshi, Kui Muraya, Fredros Okumu, Senjuti Saha, Deepika Saluja, Madhukar Pai | Australia, India, Chile, Philippines, Ghana, South Africa, Kenya, Tanzania, Bangladesh, Canada | PLOS Medicine | Journal | Commentary / Opinion piece | Power, resource and knowledge inequities in global health |
| Anti-Racism and Anti-Colonialism Praxis in Global Health-Reflection and Action for Practitioners in US Academic Medical Centres | 2021 | Zeinabou Niamé Daffe, Yodeline Guillaume and Louise C Ivers | USA | The American Society of Tropical Medicine and Hygiene | Journal | Commentary / Opinion piece | Reflective practice as a tool in decolonising global health, anti-colonialism, anti-racism |
| Applying a Power Analysis to Everything We Do: A Qualitative Inquiry to Decolonize the Global Health and Development Project Cycle | 2023 | Doreen Tuhebwe, Sarah Brittingham, Amandari Kanagaratnam, Elikem Togo, Funmilola M. OlaOlorun, Rhoda K. Wanyenze, Ndola Prata, Allysha C. Maragh-Bassb | Uganda, USA, Nigeria | Global Health: Science and Practice | Journal | Qualitative study / Literature Review | The colonial influence throughout global health project cycles, need for more equitable partnership working, redistribution of power to Global South, need for individual action and structural change. |
| Arts-based methods as a critical and decolonising process in global mental health: Reflections on popular discourse, artistic rigour and limitations | 2024 | Matthew Elliott | UK | Methods in Psychology | Journal | Qualitative study | Global mental health, masculinities and mental health, possibilities of arts-based research in decolonising mental health practices |
| Barriers to decolonising educational partnerships in global health | 2021 | John Kulesa, Nana Afua Brantuo | USA | BMJ Global Health | Journal | Literature review | Global health education, the colonial roots of global health education, equitable partnerships, decolonising health education partnerships |
| Beyond pledges: academic journals in high-income countries can do more to decolonise global health | 2021 | Bolajoko O Olusanya, Macpherson Mallewa, Felix Akpojene Ogbo | Nigeria, Malawi, Australia | BMJ Global Health | Journal | Commentary / Opinion piece | Colonial history and influence on global health, epistemic injustice, call on academic journals to promote equity, equality, diversity and inclusiveness principles related to authorship |
| Build that wall! Vaccine certificates, passes and passports, the distribution of harms and decolonial global health justice | 2021 | Gabriela Arguedas-Ramírez | Costa Rica | Journal of Global Ethics | Journal | Commentary / Opinion piece | Colonial influence on global health, decolonisation as a process, viewing the COVID-19 pandemic through a decolonial lens, ethical considerations of vaccine passports |
| Building a framework to decolonize global emergency medicine | 2023 | Monalisa Muchatuta, Shama Patel, Catalina Gonzalez Marquez, Kaushila Thilakasiri, Sreenidhi Vanyaa Manian, Jennifer Chan, Ngassa Mssika, Taryn Clark, Taylor Burkholder, Nikkole Turgeon, Vinay N Kampalath, Nivedita Poola, O Agatha Offorjebe, Adeline Dozois, Gimbo Hyuha, Oluwarotimi Vaughan-Ogunlusi, Carol McCammon, Katie Wells, Megan Rybarczk, Maria Paula Castillo, Adebisi Anthonia Adeyeye, Chris A Rees, Sanjukta Dutta, Stephanie Chow Garbern | USA, Sri Lanka, India, Tanzania, Nigeria | American Emergency Medicine Education and Training | Journal | Literature review | Colonial issues associated with emergency medicine, a path for change, the disproportionate influence of faculty and students from high-income countries (HICs) in research, academic pursuits, and instruction in global health creating one-way flow of knowledge, training opportunities, funding and funding priorities from HICs to in low and middle income countries (LMICs). |
| Can schools of global public health dismantle colonial legacies? | 2020 | Ngozi A Erondu, Dorothy Peprah, MIshal S Khan | USA, UK, Pakistan | Nature Medicine | Journal | Commentary / Opinion piece | Neo-colonial academic practice, epistemic injustice, representation in academic leadership, anti-racism at academic institutions |
| Cancer linguistics and the politics of decolonizing health communication in Coastal Tanzania: Reflections from an anthropological investigation | 2024 | Daniel W. Krugman, Athumani Litunu, Saumu Mbeya, M. Yunus Rafiq | USA, Tanzania, China | Social Science & Medicine | Journal | Commentary / Opinion piece | The construction of biomedical language and the role of language in decolonising global health, the need to dismantle incommunicability and the global order that supports it. |
| Challenges for breaking down the old colonial order in global health research: the role of research funding | 2021 | Larissa Fortunato Araújo, Flávia B Pilecco, Francisco Gustavo Silveira Correia, Marcelo José Monteiro Ferreira | Brazil | The Lancet Global Health | Journal | Commentary / Opinion piece | Inequities in the funding perpetuating colonial structures and views within global health |
| Colonialism, malaria, and the decolonization of global health | 2022 | Jesse B Bump, Ifeyinwa Aniebo | USA, Norway, Nigeria | PLOS Global Public Health | Journal | Literature review | Malaria control, colonisation, tropical medicine |
| Committing to anti-racism reforms? Three critical building blocks for global health organizations | 2022 | Mishal S Khan, Angela Obasi, Rinki Deb, Serign Jawo Ceesay | United Kingdom | PLOS Global Public Health | Journal | Commentary / Opinion piece | Anti-racism, academia, institutional racism, decolonising global health curricula, research |
| Community Engagement in Cutaneous Leishmaniasis Research in Brazil, Ethiopia, and Sri Lanka: A Decolonial Approach for Global Health | 2022 | Kay Polidano, Linda Parton, Suneth B Agampodi, Thilini C Agampodi, Binega H Haileselassie, Jayasundara M G Lalani, Clarice Mota, Helen P Price, Steffane Rodrigues, Getachew R Tafere, Leny A B Trad, Zenawi Zerihun, Lisa Dikomitis | UK, Sri Lanka, Ethiopia, Brazil | Frontiers in Public Health | Journal | Qualitative study | Community engagement, Eurocentric knowledge production in global health, colonial legacies within global health, deconstructing dominant western approaches in global health practice |
| Community First solutions for COVID-19: decolonising health crises responses | 2021 | Rachel Kiddell-Monroe, Jessica Farber, Carol Devine, James Orbinski | Canada | The Lancet Planetary health | Journal | Commentary / Opinion piece | Co-creation of responses to health crises with vulnerable communities, cultural sensitivity, self-determination, planetary health |
| Decolonising COVID-19 | 2020 | The Lancet Global Health | UK | The Lancet Global Health | Journal | Commentary / Opinion piece | COVID-19, asymmetrical power structures within global health, racism and global health practice, the colonisation of medicine, economics, and politics |
| Decolonising Global (Public) Health: from Western universalism to Global pluriversalities | 2020 | Clara Affun-Adegbulu, Opemiposi Adegbulu | Belgium, UK | BMJ Global Health | Journal | Commentary / Opinion piece | Definition and conceptualisations of decolonising global health, Eurocentric conception of global health, decolonisation at epistemic and ontological levels |
| Decolonising global health | 2024 | Julia Ngozi Chukwuma | UK | The Companion to Development Studies (4th Edition) | Book | Commentary / opinion piece | COVID-19 highlighting global health inequities. Understanding colonial roots of global health informs practice today. Issues with neo-liberal capitalism and technical (rather than system) focus approach to 'solving' global health issues and role in reinforcing power imbalances. |
| Decolonising global health by decolonising academic publishing | 2022 | Shahzad Amjad Khan | Pakistan | BMJ Global Health | Journal | Commentary / Opinion piece | Academic publishing as a place to start decolonisation, hierarchies of evidence, need for transparency |
| Decolonising global health evaluation: Synthesis from a scoping review | 2022 | Ichhya Pant, Sonal Khosla, Jasmine Tenpa Lama, Vidhya Shanker, Mohammed AlKhaldi, Aisha El-Basuoni, Beth Michel, Khalil Bitar, Ifeanyi McWilliams Nsofor | USA, Canada, UAE, Switzerland, Palestine, Nigeria | PLOS Global Public Health | Journal | Scoping review | Defining decolonising global health evaluation, identifying themes and factors associated with decolonising global health evaluation, decolonising global health as a 'journey' |
| Decolonising global health in 2021: a roadmap to move from rhetoric to reform | 2021 | Mishal Khan, Seye Abimbola, Tammam Aloudat, Emanuele Capobianco, Sarah Hawkes, Afifah Rahman-Shepherd | UK, Australia, Switzerland | BMJ Global Health | Journal | Commentary / Opinion piece | Defining decolonising global, systemic or organisational change of global health institutions, steps required to decolonise, metrics for accountability |
| Decolonising global health in the Global South by the Global South: Turning the lens inward | 2023 | Dhananjaya Sharma, Nadia Adjoa Sam-Agudu | India, Nigeria, Ghana, USA | BMJ Global Health | Journal | Commentary / opinion piece | Global health actors from the global south and their role in decolonisation (from the perspective of authors from the global south), the importance of introspection regarding colonialism / decolonising, systemic inequalities, the structural nature of coloniality, individual behaviour, knowledge creation, frameworks for taking action on decolonising global health. |
| Decolonising global health in the time of COVID-19 | 2020 | Mariam O Fofana | USA | Global Public Health | Journal | Literature review | Colonial inheritance of modern global health, COVID-19, steps for dismantling colonialism in global health |
| Decolonising global health research: Shifting power for transformative change | 2024 | Ramya Kumar, Rajat Khosla, David McCoy | Malaysia, Sri Lanka | PLOS Global Health | Journal | Commentary / opinion piece | Presentation of framework for understanding colonialism in global health research. Importance of equitable research partnerships. Call for structural change within global health research system to decolonise. Importance of transparent global health funding. |
| Decolonising global health: a Philippine perspective | 2022 | Edward Christopher Dee, Gideon Lasco | USA, Philippines | The Lancet | Journal | Commentary / Opinion piece | Rebuttal of criticism of decolonising global health, epistemic injustice, steps to decolonise, community empowerment |
| Decolonising global health: beyond ‘reformative’ roadmaps and towards decolonial thought | 2021 | Monica Mitra Chaudhuri, Laura Mkumba, Yadurshini Raveendran, Robert D Smith | Canada, USA, Switzerland | BMJ Global Health | Journal | Commentary / Opinion piece | Definitions of decolonising global health, colonialism within global health industry, need for comprehensive structural reform, necessity of reflection / introspection |
| Decolonising Global Health: Clarifying Concepts for Equitable Practice | 2024 | Emma Sophie Spanaus, Luis Eugenio De Souza | Switzerland, Germany, Brazil | Epidemiology & Public Health | Journal | Qualitative study / Literature Review | Power dynamics, importance of traditional and community-based knowledge, structural determinants of colonialism in global health, need for equitable partnerships, reform of global health curriculums |
| Decolonising global health: if not now, when? | 2020 | Ali Murad Büyüm, Cordelia Kenney, Andrea Koris, Laura Mkumba, Yadurshini Raveendran | USA | BMJ Global Health | Journal | Commentary / Opinion piece | Colonial structure of global / public health, individual duty, actions to decolonise global health, COVID-19, reflective practice, paradigm / leadership / knowledge shift |
| Decolonising global health: transnational research partnerships under the spotlight | 2020 | David S Lawrence, Lioba A Hirsch | UK, Botswana | International health | Journal | Literature review | Clinical trials, transnational research partnerships, decolonising global health research |
| Decolonising global health: where are the Southern voices? | 2021 | Samuel Oti, Jabulani Ncayiyana | Kenya, South Africa | BMJ Global Health | Journal | Commentary / Opinion piece | Definitions of decolonising global health, need for pragmatism over further theorising, decolonising from perspective of Global South |
| Decolonising global health: why the new Pandemic Agreement should have included the principle of subsidiarity | 2024 | Thana C de Campos-Rudinsky, Sarah L Bosha, Daniel Wainstock, Sharifah Sekalala, Sridhar Venkatapuram, Caesar Alimsinya Atuire | Chile, USA, Brazil, UK, South Africa, Ghana | The Lancet Global Health | Journal | Commentary / opinion piece | Pandemic Agreement, subsidiarity, recommendations for global health governance, engaging local and community level actors in global health policy and mechanisms |
| Decolonising ideas of healing in medical education | 2020 | Amali U. Lokugamage, Tharanika Ahillan, S. D. C. Pathberiya | UK | Journal of Medical Ethics | Journal | Literature review | Medical education, enlightenment, healing, indigenous knowledge, colonial history of global health |
| Decolonising integrative health: learning more from and elevating the voices of rich but often neglected health traditions | 2022 | Jon Wardle | Australia | Advances in Integrative Medicine | Journal | Commentary / Opinion piece | Decolonising integrative health and traditional medicine |
| Decolonising 'man', resituating pandemic: an intervention in the pathogenesis of colonial capitalism | 2021 | Rosemary J Jolly | USA | Medical Humanities | Journal | Commentary / Opinion piece | Decolonial pandemic history, individual and institutional practice for decolonisation |
| Decolonising public and planetary health, or Chthulecene mediations | 2023 | Jan Gresil Kahambing | China | Journal of Public Health | Journal | Commentary / Opinion piece | Structural inequalities in global health, funding, power asymmetries between actors from high and low income countries working in global health. |
| Decolonising qualitative research to explore the experiences of Manitoba’s urban Indigenous population living with type 2 diabetes mellitus, obesity and bariatric surgery | 2020 | Krista Hardy, Kathleen Clouston, Marta Zmudzinski, Melinda Fowler-Woods, Geraldine Shingoose, Amanda Fowler-Woods, Felicia Daeninck, Andrew Hatala, Ashley Vergis | Canada | BMJ Open | Journal | Qualitative study | Decolonising qualitative research in practice, cultural and community engagement |
| Decolonising violence against women research: a study design for co-developing violence prevention interventions with communities in low and middle income countries (LMICS) | 2021 | Jenevieve Mannell, Safua Akeli Amaama, Ramona Boodoosingh, Laura Brown, Maria Calderon, Esther Cowley-Malcolm, Hattie Lowe, Angélica Motta, Geordan Shannon, Helen Tanielu, Carla Cortez Vergara | UK, New Zealand, Samoa, Peru | BMC Public Health | Journal | Case study | Decolonising violence against women research, power dynamics in western research practice, community participation and co-design |
| Decolonization and antiracism: intersecting pathways to global health equity | 2024 | Collins O. Airhihenbuwa, Chandra Ford, Juliet Iwelunmor, Derek M. Griffith, Khadijah Ameen, Teri Murray, Ucheoma Nwaozuru | USA | Ethnicity & Health | Journal | Commentary / opinion piece | Decolonisation and antiracism, colonisation, health equity, public health critical race praxis, global structural racism |
| Decolonization in health professions education: reflections on teaching through a transgressive pedagogy | 2016 | Ruth Rodney | Canada | Canadian Medical Education Journal | Journal | Qualitative study | Transgression in medical education that can lead to decolonising, acknowledging the HIC educators' positionality in LMICs, reflective practice, the historical colonial roots of global health |
| Decolonizing Epidemiological Research: A Critical Perspective | 2023 | Yusuff Adebayo Adebisi | UK | Avicenna Journal of Medicine | Journal | Commentary / opinion piece | Important role for indigenous and marginalized populations, power imbalances in epidemiological research, distribution of research funding, authorship in epidemiological research, journal editorship. |
| Decolonizing global health — what does it mean for us? | 2023 | Birger C. Forsberg, Jesper Sundewall | Sweden, South Africa | European Journal of Public Health | Journal | Commentary / Opinion piece | Mapping flow of funds in global health, research on use and efficiency of funding in global health, twinning of research programmes for HIC and LMIC based institutions. |
| Decolonizing global health by engineering equitable relationships | 2024 | Nature Reviews Bioengineering | Unspecified | Nature Reviews Bioengineering | Journal | Commentary / opinion piece | Bioengineering; equitable relationships to drive innovation; hierarchical assumptions and disregard/lack of understanding of local knowledge and contexts. |
| Decolonizing global health curriculum: from fad to foundation | 2023 | Anna Kalbarczyk, Sylvie Perkins, Sabreena N. Robinson, Mahnoor K. Ahmed | USA | Frontiers in Education | Journal | Qualitative study | Interpretation of decolonising global health in educational programmes, guidance, protected time and support for decolonising activities, |
| Decolonizing global health education: rethinking institutional partnerships and approaches | 2021 | Quentin G Eichbaum, Lisa V Adams, Jessica Evert, Ming-Jung Ho, Innocent A Semali, Susan C van Schalkwyk | USA, Tanzania, South Africa | Academic Medicine: Journal of the Association of American Medical Colleges | Journal | Commentary / Opinion piece | Intersectionality of colonialism, power dynamics and neo-colonialist assumptions, approaches to challenge colonial paradigms in global health |
| Decolonizing global health from the perspectives of global health actors in Low-middle Income Countries | 2021 | Sedem Adiabu | USA | Emory University Theses and Dissertations | Grey literature | Qualitative study | Interpretations of decolonising global health, political manipulation, hypocrisy, distrust, partnership working, steps needed to decolonise |
| Decolonizing Global Health Research: Perspectives from US and International Global Health Trainees | 2023 | Matthew DeCamp, Limbanazo Matandika, Lameck Chinula, Jorge L Cañari-Casaño, C Hunter Davis, Emily Anderson, Marlena McClellan, Benjamin H Chi, Valerie A Paz-Soldan | USA, Malawi, Peru | Annals of Global Health | Journal | Mixed methods study | Exploration of research fellow's perceptions of decolonising, definitions, awareness and attitudes to decolonising global health, |
| Decolonizing Global Health: A critical perspective from Latin America | 2024 | Vivian Laurens, César Abadía-Barrero | USA | The Routledge Handbook of Anthropology and Global Health | Book | Commentary / opinion piece | Decolonising global health from the perspective of Latin American Social Medicine, disconnect between global health funders / leaders and local knowledge and understanding of needs. Neo-liberalisation of global health, emphasis on individual agency. Indigenous concept and practice of 'Buen Vivir' in rural communities in Colombia as an example of decolonial approach in action. |
| Decolonizing Global Health: A Moment to Reflect on A Movement | 2021 | Madhukar Pai | Canada | Forbes | Grey literature | Commentary / Opinion piece | COVID-19, the plurality of decolonising global health, need for structural change, awareness of decolonising global health in the Global South, need to understand the history and meaning of decolonising global health |
| Decolonizing Global Health: Increasing Capacity of Community Health Worker Programs | 2023 | Pamela Avila | USA | Annals of Global Health | Journal | Commentary / opinion piece | Community health workers, the ethnicity of global health volunteer experiences and researchers, sustainability |
| Decolonizing global health: what should be the target of this movement and where does it lead us? | 2022 | Xiaoxiao Kwete, Kun Tang, Lucy Chen, Ran Ren, Qi Chen, Zhenru Wu, Yi Cai, Hao Li | China | Global Health Research and Policy | Journal | Commentary / Opinion piece | Historical colonial context and its ongoing influence in global health, meaning of decolonising global health, systemic change, the development of decolonial practice |
| Decolonizing healthcare innovation: Low cost solutions from low-income countries | 2024 | Matthew Harris | UK | Decolonizing healthcare innovation: Low cost solutions from low-income countries | Book | Commentary / Opinion piece | Global health, innovation, decolonisation through the promotion of knowledge, expertise and innovation from contexts that have previously been neglected, HICs learning from and making use of innovation from LMICs. |
| Decolonizing Indigenous health: Generating a productive dialogue to eliminate Rheumatic Heart Disease in Australia | 2021 | Emma Haynes, Roz Walker, Alice G. Mitchell, Judy Katzenellenbogen, Heather D’Antoine, Dawn Bessarab | Australia | Social Science and Medicine | Journal | Qualitative study | Importance of cultural safety and reflexivity on power and privilege, systemic change, combining indigenous knowledge with biomedical and social science approaches |
| Decolonizing the decolonization movement in global health: A perspective from global surgery | 2022 | Emmanuel Bua, Saad Liaqat Sahi | Uganda, USA | Frontiers in Education | Journal | Commentary / opinion piece | Discussion on decolonising global health by an author from a low and middle income country. Decolonisation of global surgery, barriers to health education in global surgery, issues with surgical missions to LMICs, knowledge sharing and bilateral exchange |
| Developing an agenda for the decolonization of global health | 2024 | David McCoy, Anuj Kapilashrami, Ramya Kumar, Emma Rhule, Rajat Khosla | Malaysia, England, Sri Lanka | Bulletin of the World Health Organization | Journal | Commentary / opinion piece | Structural nature of power inequities within global health, importance of finance, the commodification of healthcare (demonstrated by profits made through COVID-19 response), social medicine as anticolonial tradition in Latin America, need for stronger role for WHO, more inclusive and diverse global health governance. |
| Dialogical reflexivity towards collective action to transform global health | 2021 | Harvy Joy Liwanag, Emma Rhule | Malaysia | BMJ Global Health | Journal | Commentary / Opinion piece | Reflexivity and what reflexivity requires in order to contribute to decolonising global health, building cultures of reflexivity, actions to decolonise global health |
| Diversifying Implementation Science: A Global Perspective | 2022 | Sophia M. Bartels, Shabab Haider, Caitlin R. Williams, Yameen Mazumder, Latifat Ibisomi, Olakunle Alonge, Sally Theobald, Till Bärnighausen, Juanita Vasquez Escallon, Mahnaz Vahedi, Rohit Ramaswamy, Malabika Sarker | USA, South Africa, Nigeria, UK, Germany, Switzerland, Bangladesh | Global Health: Science and Practice | Journal | Commentary / Opinion piece | Role of leadership in increasing the influence of researchers and practitioners from LMICs, equity in implementation research, community participation, global action |
| Do we need to decolonise global health? | 2021 | Tammam Aloudat | Switzerland | Action to Decolonise Global Health | Grey literature | Commentary / Opinion piece | Colonial legacies in global health, inequitable power structures, disparities in health outcomes, action to decolonise global health |
| Efforts to a Belief and Decolonize Global Health | 2023 | Salim Omambia Matagi | Kenya | Journal of Advances in Medicine and Medical Research | Journal | Qualitative study / Literature Review | Decolonisation of academic publishing and research, global health as a colonial tool, individual duty of global health actors to drive move away from colonial model of global health. |
| Eliminating the White Supremacy Mindset from Global Health Education | 2022 | Agnes Binagwaho, Brianna Ngarambe, Kedest Mathewos | Rwanda | Annals of Global Health | Journal | Commentary / Opinion piece | Roots of colonisation, white supremacy, acknowledgement of expertise within LMICs, epistemic injustice |
| Epistemic injustice in academic global health | 2021 | Himani Bhakuni, Seye Abimbola | Netherlands, USA | The Lancet Global health | Journal | Commentary / Opinion piece | Epistemic injustice in global health, research, academia |
| Forming a coalition of the willing to decolonise global health. Is it possible, what impact could it have, and what next? | 2021 | Development Reimagined | China, Kenya | Development Reimagined | Grey literature | Commentary / Opinion piece | Practical steps / a framework for decolonising global health, inequitable power dynamics, institutional scepticism about decolonising global health, lessons from COVID-19 |
| Global Health and Decolonisation in Higher Education: Examining the Attitudes, Perceptions and Possibilities of Educators | 2023 | Amani Eltayb, Karin Båge, Abdalla Mohamed Ibrahim, Natalie Jellinek, Raman Preet, Zoe Säflund, Jennifer Valcke | Sweden, Somalia | Philosophy and Theory in Higher Education | Journal | Mixed methods study | Exploring perceptions and meaning of decolonisation amongst global health educators, value and relevance of decolonisation, decolonising and the relationship with global power structures. |
| Global health and the elite capture of decolonization: On reformism and the possibilities of alternate paths | 2023 | Daniel W. Krugman | USA | PLOS Global Public Health | Journal | Literature review | Radical decolonial theory, social change, the "buzzwordification" of decolonising global health, the elite capture of decolonisation. |
| Global health education in high-income countries: confronting coloniality and power asymmetry | 2022 | Hoda Sayegh, Christina Harden, Hijab Khan, Madhukar Pai, Quentin G Eichbaum, Charles Ibingira, Gelila Goba | USA, Canada, Uganda | BMJ Global Health | Journal | Commentary / Opinion piece | Global health education, the changes needed to address power asymmetries in global health education |
| Global Health Partnerships and the Brocher Declaration: Principles for Ethical Short-Term Engagements in Global Health | 2022 | Shailendra Prasad, Myron Aldrink, Bruce Compton, Judy Lasker, Peter Donkor, David Weakliam, Virginia Rowthorn, Efua Mantey, Keith Martin, Francis Omaswa, Habib Benzian, Erwin Clagua-Guerra, Emilly Maractho, Kwame Agyire-Tettey, Nigel Crisp, Ramaswami Balasubramaniam | USA, Ghana, Ireland, Guatemala, Uganda, UK, India | Annals of Global Health | Journal | Commentary / Opinion piece | Short terms experiences in global health (HIC to LMIC) as a symptom of a colonising mindset in global health, COVID as a break and chance to recalibrate, individual duty in enacting change |
| Global Health: Reimagining Perspectives | 2020 | Fernando De Maio, Jonatan Konfino | USA, Argentina | Health Disparities in Allergic Diseases | Book | Commentary / Opinion piece | Human rights, epidemiology, collaboration and partnership working |
| Global mental health research and practice: a decolonial approach | 2022 | Eliut Rivera-Segarra, Franco Mascayano, Lubna Alnasser, Els van der Ven, Gonzalo Martínez-Alés, Sol Durand-Arias, Maria Francesca Moro, Elie Karam, Ruthmarie Hernández-Torres, Sebastián Alarcón, Alíxida Ramos-Pibernus, Rubén Alvarado, Ezra Susser | Puerto Rico, USA, Saudi Arabia, Netherlands, Spain, Mexico, Italy, Lebanon, Chile | The Lancet Psychiatry | Journal | Commentary / Opinion piece | The reproduction of power imbalances and patterns of oppression through global mental health, the need for individual action and active resistance, access to conversations on decolonising, epistemic justice, pragmatic solidarity and sovereign acts as routes to decolonising global mental health |
| Has Authorship in the Decolonizing Global Health Movement Been Colonized? | 2023 | Chris A. Rees, Gouri Rajesh, Hussein K. Manji, Catherine Shari, Rodrick Kisenge, Elizabeth M. Keating, Ikechukwu U Ogbuanu, Kitiezo Aggrey Igunza, Richard Omore, Karim P. Manji | USA, Tanzania, Sierra Leone, Kenya | Annals of Global Health | Journal | Qualitative study / Literature Review | Colonisation of authorship, understandings of decolonising in high and low and middle income countries, the need for representation from authors from low- and middle-income countries. |
| How and Why Should We Decolonize Global Health Education and Research? | 2022 | Ipek Demir | UK | Annals of Global Health | Journal | Commentary / Opinion piece | Health education, research, racism, the potential for decolonised global health curricula to be more rigorous and deliver better health outcomes, legacies of coloniality in health outcomes, actions for decolonising global health education |
| In the wake: Interpreting care and global health through Black geographies | 2019 | Lioba A Hirsch | UK | Area | Journal | Commentary / Opinion piece | Critical theoretical reflections on global health interventions in postcolonial societies |
| Is decolonisation sufficient? | 2022 | Sana Qais Contractor, Jashodhara Dasgupta | Belgium, Norway, India | BMJ Global Health | Journal | Commentary / opinion piece | Local and global power hierarches, intersectionality, the types of actors within global health, positionality. |
| Is it possible to decolonise global health institutions? | 2021 | Lioba A Hirsch | UK | The Art of Medicine | Journal | Commentary / Opinion piece | Structural determinants of power, structural violence and hurt inflicted by global health institutions, need for action rather than tokenistic efforts to decolonise |
| It’s Time to Decolonize the Decolonization Movement | 2021 | Ijeoma Nnodim Opara | USA | PLOS Blogs: Speaking of Medicine and Health | Grey literature | Commentary / Opinion piece | Power asymmetries, critical analyses of global health, who is currently benefitting from decolonising movement, steps needed to decolonise global health |
| Knowledge is Power: Assessing Academic Decolonization through Bidirectionality of Training in Global Health Fellowships | 2022 | Rebecca Fujimara, Yalda Jabbarpour | USA | McGill Journal of Global Health | Journal | Literature review | The idea of 'bi-directional power', power imbalances between global north and south, the benefits of global health fellowships for those from high income countries. |
| Letter to the Editor: Leveraging ChatGPT to democratize and decolonize global surgery: Large language models for small healthcare budgets | 2024 | Hinpetch Daungsupawong, Viroj Wiwanitkit | Laos, India, Nigeria | World Journal of Surgery | Journal | Commentary / opinion piece |  |
| Missing in action: a scoping review of gender as the overlooked component in decolonial discourses | 2024 | Tiffany Nassiri-Ansari, Emma Louise Margaret Rhule | Malaysia | BMJ Global Health | Journal | Qualitative study / Literature Review | Gender as an overlooked dimension within decolonising global health literature, intersectionality, colonial impact on concepts of identity and knowledge, the need for the integration of a gender lens within work to decolonise global health. |
| Offline: The case for global health | 2023 | Richard Horton | UK | The Lancet | Journal | Commentary / Opinion piece | Origins of global health, link between funding and influence, scientific publishing and access to journals, structural racism. |
| Offline: The myth of "decolonising global health" | 2021 | Richard Horton | UK | The Lancet | Journal | Commentary / Opinion piece | Neo-colonialism, the structural determinants of colonialism in global health, decolonising global health as a challenge, the need for action over rhetoric |
| One Health: A social science discussion of a global agenda | 2022 | Jean Estebanez, Pascal Boireau | France | Parasite | Journal | Commentary / Opinion piece | One Health and its roots in the colonial period, re-legitimisation of local and non-human / animal knowledge, decolonisation of One Health to prevent epidemic emergence, |
| Post-decolonisation: Global Health and Global Surgery's Coming of Age | 2022 | Bhavna Chawla, Judith Lindert, Dhananjaya Sharma, | Switzerland, Germany, India | The Indian Journal of Surgery | Journal | Commentary / Opinion piece | Global surgery, medical education, disconnect between global north and the 'beneficiaries' of global health, post-colonial mindset of those working in global health |
| Powerful ideas? Decolonisation and the future of global health | 2022 | Mark Hellowell, Patricia Nayna Schwerdtle | UK, Germany | BMJ Global Health | Journal | Commentary / Opinion piece | Knowledge, universalism, colonial origins of global health, asymmetries in the distribution of epistemic authority and decision making power in global health, structural determinants of power within global health, global health's Eurocentric conception of humanity, COVID-19 response |
| Prioritizing equity and inclusion in global health dermatology | 2021 | Aileen Y Chang, Miriam Laker-Oketta, Sarah J. Coates | USA, Uganda | International Journal of Women's Dermatology | Journal | Commentary / Opinion piece | Dermatology, medical education, actions to decolonize global health dermatology through research, training and service delivery, funding, epistemic injustice, representation, the burden practitioners and researchers from HICs place on the LMIC settings in which they may practice. |
| Radiologists’ Role in Decolonizing Global Health | 2024 | Farouk Dako, Toma S. Omofoye, John Scheel | USA | Journal of the American College of Radiology | Journal | Commentary / opinion piece | Radiology and decolonising global health, capacity strengthening, inequitable access to technology, brain drain, parachute research. |
| Realist evaluation in times of decolonising global health | 2022 | Dimitri Renmans, Nandini Sarkar, Sara Van Belle, Clara Affun-Adegbulu, Bruno Marchal, Ferdinand C Mukumbang | Belgium, USA | The International Journal of Health Planning and Management | Journal | Commentary / Opinion piece | Role of realistic evaluation in decolonising global health, importance of engaging with power inequities in global health, overreliance on Western-based theories and knowledge, participatory research methods. |
| Reflections on ‘Decolonizing’ Big Data in Global Health | 2022 | Danya M Qato | USA | Annals of Global Health | Journal | Commentary / Opinion piece | The principles of decolonising global health should be applied to the collection and use of data in global health, given its important role in global health, COVID-19, leadership by people who 'populate the dataset', need for decolonising agenda to be clear and understandable, ownership of data |
| Reimagining global health: From decolonisation to indigenization | 2021 | Suzanne Hindmarch, Sean Hillier | Canada | Global Public Health | Journal | Commentary / Opinion piece | Decolonisation, indigenous ontologies and health expertise, dominance of western principles and ideologies in global health |
| Rethinking development interventions through the lens of decoloniality in sub-Saharan Africa: The case of global health | 2022 | L Gautier, Y Karambe, J-P Dossou, O M Samb | Canada, Mali, Benin, Belgium | Global Public Health | Journal | Commentary / Opinion piece | Western dominance of global health, approaches to decolonising global health, the inability of global health to engage with the 'structures and mental models' of African populations. |
| Shifting Power in Global Health - Decolonising discourses - Series Synthesis | 2022 | Tiffany Nassiri-Ansari, Emma Rhule | Malaysia | Summary of United National University, Development Reimagined, and Wilton Park Virtual Dialogue Series | Grey Literature | Commentary / opinion piece | Exploration of decoloniality through language, representation, and positionality, funding, power dynamics, agency and equity as tools to reimagine collective development. |
| Swampscott in International Context: Expanding Our Ecology of Knowledge | 2016 | Christopher C. Sonn | Australia | American Journal of Community Psychology | Journal | Commentary / Opinion piece | Colonisation of knowledge, the 'dynamics of dominance and marginality in knowledge production in psychology', opportunities to decolonise community psychology. |
| Talk the talk and walk the walk: a novel training for medical students to promote decoloniality in global health | 2024 | Leah Ratner, Shela Sridhar, Sheila Owusu, Samantha L. Rosman, Rose L. Molina, Jennifer Kasper | USA, Ghana | Frontiers in Education | Journal | Mixed methods study | Decoloniality within medical education, moving from theory to practical application of decolonial principles, need for accountability and collective action, need for institutional and structural as well as individual-level change. |
| Talking Points: Catherine Kyobutungi on global health | 2022 | Helga Groll | UK | eLife | Grey literature | Commentary / Opinion piece | Helicopter models of research, mismatch between the priorities of funders and those based in Africa, need to move from discussion to action to decolonise. |
| The ‘decolonization of global health’ agenda in Africa: harnessing synergies with the continent’s strategic aspirations | 2023 | Aloysius Ssennyonjo, Phillip Wanduru, Elizabeth Omoluabi, Peter Waiswa | Uganda, Belgium, Sweden, Senegal, South Africa | European Journal of Public Health | Journal | Commentary / opinion piece | Call for Africa-based development practitioners to 'decolonise from within', need for egalitarian partnership working, need for African-led, African based models for capacity strengthening. |
| The activists trying to 'decolonize' global health | 2019 | Andrew Green | Germany | DevEX | Grey literature | Commentary / Opinion piece | Activism, race and political economy in the delivery of health services, structural determinants of global health, decolonial theory |
| The cognitive dissonance discourse of evolving terminology from colonial medicine to global health and inaction towards equity - A Preventive Medicine Golden Jubilee Article | 2022 | Delivette Castor, Luisa N Borrell | USA | Preventive medicine | Journal | Qualitative study | Language, equity / inequity, social justice, decolonising global health terminology |
| The feudal structure of global health and its implications for decolonisation | 2022 | Vikash Ranjan Keshri, Soumyadeep Bhaumik | Australia, India | BMJ Global Health | Journal | Commentary / Opinion piece | Euro-American hegemony over knowledge ecosystems in global health is supported by global health structures similar to old feudal power structures from colonial era, the need to dismantle structure of global health to move decolonising global health forward, parachuting-in of skills from high income to low and middle income countries, the colonial historical roots of global and international health |
| The foreign gaze: authorship in academic global health | 2019 | Seye Abimbola | Australia | BMJ Global Health | Journal | Commentary / Opinion piece | Entrenched power asymmetries in global health partnerships, the 'foreign gaze' in global health and its consequences and influence. |
| The future of global health education: training for equity in global health | 2016 | Lisa V. Adams, Claire M. Wagner, Cameron T. Nutt, Agnes Binagwaho | USA, Switzerland, Rwanda | BMC Medical Education | Journal | Literature review | Need for partnership working between the global north and global south, traditional (colonial) roles institutions from the global north and south play, models for equitable training partnerships in global health |
| The Rhetoric of Decolonizing Global Health Fails to Address the Reality of Settler Colonialism: Gaza as a Case in Point | 2024 | Eivind Engebretsen, Mona Baker | Norway, UK | International Journal of Health Policy and Management | Journal | Commentary / Opinion piece | Decolonising knowledge translation, colonial structures of health knowledge system, need for action, reflection and theorisation, need for clear definitions of decolonising global health. |
| The rise of non-communicable disease (NCDs) in Mozambique: decolonising gender and global health | 2021 | Claire Somerville, Khatia Rebeca Munguambe | Mozambique, Switzerland | Gender & Development | Journal | Qualitative study | Non-communicable disease, gender, healthcare systems, Mozambique, dominance of global North in setting global health agenda, structural inequalities in global health, biomedical explanatory models of illness and their ‘fit’ with existing or local models of illness and diagnosis. |
| The truth about decolonising global health worth spreading | 2022 | Juliet Iwelunmor | USA | The Lancet | Journal | Commentary / Opinion piece | Decolonising global health will take a long time, the process of decolonising, struggle, decolonising as an 'unconventional movement' |
| The way forward in decolonising global health | 2023 | Sudeep Adhikari, Irene Torres, Elizabeth Oele | Nepal, Ecuador, Kenya | The Lancet Global Health | Journal | Commentary / opinion piece | Recommendations for decolonising global health; reduce migration of health workers from LMICs, globalise clinical trials. |
| The words we choose matter: recognising the importance of language in decolonising global health | 2021 | Franziska Hommes, Helena Brazal Monzó, Rashida Abbas Ferrand, Meggan Harris, Lioba A Hirsch, Emilie Koum Besson, John Manton, Toyin Togun, Robindra Basu Roy | UK, Germany, Zimbabwe, Spain, Gambia | The Lancet Global Health | Journal | Commentary / Opinion piece | Importance of language and the terminology used to discuss global health, publishing in English and difficulties with translation, Anglocentrism as a barrier for readers, authors and researchers, the historical context of domination, dependence, and subordination in global health |
| Theories, models and best practices for decolonizing global health through experiential learning | 2023 | Steven R. Hawkes, Jenna L. Hawks, Heather S. Sullivan | USA | Frontiers in Education | Journal | Literature review | Experiential and transformative learning, critical reflection, educational practice, collaboration with local populations and community partners. |
| Theory from the South: a decolonial approach to the psychology of global inequality | 2017 | Glenn Adams, Sara Estrada-Villalta | USA | Current Opinion in Psychology | Journal | Commentary / Opinion piece | Coloniality within global health and 'modernity and modern mentalities', global security |
| Thinking the post-colonial in medical education | 2008 | Alan Bleakley, Julie Brice, John Bligh | UK | Medical Education | Journal | Commentary / Opinion piece | Post-colonial theory and medical education, the western medical curriculum's influence globally and its influence on alternative or local approaches to medicine and medical education |
| To Decolonize Global Health, We Must Examine the Global Political Economy | 2021 | Pooja Yerramilli | USA, India | Think Global Health | Grey literature | Commentary / Opinion piece | COVID-19, international aid, the commercial determinants of health, public sector spending, the prioritisation of financial sustainability over access to health care |
| To decolonize global surgery and global health we must be radically intentional | 2023 | Denis A. Foretia | Cameroon | The American Journal of Surgery | Journal | Commentary / Opinion piece | Medical tourism, health system strengthening, power dynamics within clinical teams, saviourism. |
| Toward a decolonized healthcare ethics: Colonial legacies and the Siamese crocodile | 2020 | Luis Cordeiro-Rodrigues | China | Developing world bioethics | Journal | Commentary / Opinion piece | COVID-19, interdependency, colonial legacy of hostility, global distribution of resources to support healthcare |
| Towards attainment of Indigenous health through empowerment: resetting health systems, services and provider approaches | 2021 | Cheryl Barnabe | Canada | BMJ Global Health | Journal | Case study | Colonial policies and their implications for the health of indigenous populations of commonwealth countries, imposition of models of healthcare without regard to context or indigenous knowledge, community led healthcare delivery |
| Transcending the guilt of global health | 2019 | Richard Horton | UK | Lancet | Journal | Commentary / Opinion piece | Academic institutions and their colonial legacies, barriers to academic publication for those from low-income and middle-income countries, epistemic injustice, the power of journals and their editorial staff |
| Transformational learning to decolonise global health | 2021 | Amali U Lokugamage, Sarah H M Wong, Nathan M A Robinson, Sithira D C Pathberiya | UK | The Lancet | Journal | Commentary / Opinion piece | Colonial era power hierarchies in global health, prerequisites for systemic change, programmes of transformational learning to decolonise global health |
| Transforming global health through equity-driven funding | 2021 | Jacob O Olusanya, Olufunmilayo I Ubogu, Fidelis O Njokanma, Bolajoko O Olusanya | Nigeria, South Africa | Nature Medicine | Journal | Commentary / Opinion piece | The philosophy of global health reinforces inequalities and promotes the interests of the Global North, funding structures and flows do not reflect equality, equity, diversity and inclusion (EEDI) principles, the steps that funders can take to decolonise global health |
| Transforming global health: decoloniality and the human condition | 2024 | Raphael Lencucha | Canada | BMJ Global Health | Journal | Commentary / opinion piece | Risk of the 'totalising gaze' by focussing solely on decolonising without recognition of complex human behaviour e.g. tendency to seek power, emphasise difference. Structural and epistemic inequalities in global health. |
| Undoing supremacy in global health will require more than decolonisation | 2020 | Keerti Gedela | UK | The Lancet | Journal | Commentary / Opinion piece | Global health architecture shaped by supremacy and colonisation, LMIC internal power relations, personal interest and systemic change |
| Undoing supremacy in global health will require more than decolonisation - Authors' reply | 2021 | Seye Abimbola, Madhukar Pai | Australia, Canada | Lancet | Journal | Commentary / Opinion piece | Colonialism and power asymmetry between high-income countries and low-income and middle-income countries, lessons from COVID-19, research practice |
| Unequal ecosystems of global health authorial expertise: Decolonising noncommunicable disease | 2021 | Clare Herrick, Oritsematosan Okpako, James D.A. Millington | UK | Health & Place | Journal | Literature review | Authorship on papers written about non-communicable diseases, the geographies of authorial expertise and its reproduction of the inequities of coloniality. |
| We Need to Decolonize | 2022 | Barry, Michele | USA | Stanford Centre for Innovation in Global Health: Global Health Spotlight Newsletter | Grey literature | Commentary / Opinion piece | Legacy of imperial power, inequitable nature of partnerships in global health between Global North and South, structural inequities and links to poorer outcomes, need to dismantle imperial power structures, need for self-scrutiny and openness to change. |
| What Do Global Health Practitioners Think about Decolonizing Global Health? | 2022 | Madelon L Finkel, Marleen Temmermann, Fatima Suleman, Michele Barry, Melissa Salm, Agnes Bingawaho, Peter H Kilmarx | USA, Kenya, South Africa, Rwanda | Annals of Global Health | Journal | Literature review | Perceptions of equity and power imbalances in global health partnerships, leadership, funding, authorship and academic initiatives as target areas for decolonisation |
| What Is Global Health: Science and Practice Doing to Address Power Imbalances in Publishing? | 2020 | Sonia Abraham, Stephen Hodgins, Abdulmumin Saad, Madeleine Short Fabicc | USA, Canada | Global Health: Science and Practice | Journal | Commentary / Opinion piece | Decolonising academic publication, epistemic injustice, local of power structures located in high-income countries while its implementation is located in LMICs, colonial history, funding, and social and economic structures, asymmetries of power and expertise, the influence of COVID-19. |
| What research evidence can support the decolonisation of global health? Making space for deeper scholarship in global health journals | 2023 | Sudha Ramani, Eleanor Beth Whyle, Nancy Kagwanja | India, South Africa, Kenya | Lancet Global Health | Journal | Commentary / opinion piece | Decolonising research practice, the important of local context and the importance of including experiential and local knowledge in global health interventions, need to challenge power imbalances and hegemonic discourses, systemic change in global health, importance of moving beyond biomedical approach and primacy of clinical trials or observational studies to incorporate qualitative and interpretive methods. |
| Why ‘elevating country voice’ is not decolonizing global health: A frame analysis of in-depth interviews | 2023 | Michael Kunnuji , Yusra Ribhi Shawar, Rachel Neill, Malvikha Manoj, Jeremy Shiffman | Nigeria, USA, | PLOS Global Health | Journal | Qualitative study / Literature Review | Decolonising global health's focus on addressing legacy of colonialism, neoliberal exploitation, emphasis on systemic change. Feasibility and desirability of wholesale decolonisation of global health. |
| Why and for whom are we decolonising global health? | 2021 | Ong’era F Mogaka, Jenell Stewart, Elizabeth Bukusi | Kenya, USA | The Lancet Global Health | Journal | Commentary / Opinion piece | Donor programmes, shift to local ownership, need for radical transformation and systematic change, white supremacy and the vested interests of established global health practitioners in high income countries, structural determinants of inequities within global health |
| Why It's important for future healthcare professionals to understand 'decolonizing global health', and how this can be done | 2020 | Nardin Farag | Canada | McGill University Perspectives Blog | Grey literature | Commentary / Opinion piece | Conceptualisation and understanding of decolonising global health, structural determinants of power in global health, need for reflective practice in global health, the repercussions of colonial history within global health |
| Will global health survive its decolonisation? | 2020 | Seye Abimbola, Madhukar Pai | Australia, Canada | The Lancet | Journal | Commentary / Opinion piece | Colonial roots of global health, representation within global health leadership, equitable and just global health architecture and how that compares to global health structures currently in place. |
