## Supplementary material for "Decolonising Global Health: a scoping review": S1 Appendix - Search Strategy

**S1 Appendix: Decolonising global health: A scoping review search strategy**

The following pages set out the search strategy used for the scoping review. We ran two searches, the first in September 2022, and an updated search in August 2024.

**Search one: September 2022**

**MEDLINE(R) ALL 1946 to September 08, 2022**

| **#** | **Searches** | **Results** |
| --- | --- | --- |
| 1 | Global health.tw,kw. | 33352 |
| 2 | Global public health.tw,kw. | 8488 |
| 3 | global mental health.tw,kw. | 1138 |
| 4 | Transnational partnership*.tw,kw. | 7 |
| 5 | International partnership*.tw,kw. | 385 |
| 6 | World health.tw,kw. | 80615 |
| 7 | (Transnational cooperat* or transnational co-operat*).tw,kw. | 26 |
| 8 | transnational collaborat*.tw,kw. | 32 |
| 9 | International health.tw,kw. | 3925 |
| 10 | (International coopperat* or international co operat*).tw,kw. | 234 |
| 11 | international collaborat*.tw,kw. | 7356 |
| 12 | (Partnership* adj7 (LMIC* or HIC* or income countr*)).tw,kw. | 165 |
| 13 | *Global Health/ | 22216 |
| 14 | *Internationality/ or *International Cooperation/ or *International Educational Exchange/ | 28363 |
| 15 | or/1-14 | 170273 |
| 16 | Decoloni*.tw,kw. | 1949 |
| 17 | (Racism or racist).tw,kw. | 7389 |
| 18 | ((race* or racial*) adj3 (prejudic* or bias* or discriminat*)).tw,kw. | 3965 |
| 19 | (Oppress* adj5 (minorit* or people of colo?r or person* of colo?r or indigenous)).tw,kw. | 92 |
| 20 | (Oppress* adj5 (racial* or race*)).tw,kw. | 130 |
| 21 | (privilege* adj5 (racial* or race*)).tw,kw. | 97 |
| 22 | Whiteness.tw,kw. | 1158 |
| 23 | White privilege*.tw,kw. | 112 |
| 24 | white suprem*.tw,kw. | 166 |
| 25 | (white adj2 majorit*).tw,kw. | 578 |
| 26 | colonial*.tw,kw. | 7267 |
| 27 | post-colonial*.tw,kw. | 236 |
| 28 | (Historical coloni* or historical coloni*).tw,kw. | 53 |
| 29 | ((high* income* countr* or HIC) adj3 domina*).tw,kw. | 16 |
| 30 | Northern ventriloquis*.tw,kw. | 1 |
| 31 | Epistem* violence.tw,kw. | 21 |
| 32 | Epistem* oppress*.tw,kw. | 5 |
| 33 | Epistem* injustice*.tw,kw. | 122 |
| 34 | epistem* knowledge*.tw,kw. | 18 |
| 35 | western epistem*.tw,kw. | 12 |
| 36 | indigenous epistem*.tw,kw. | 22 |
| 37 | Southern voice*.tw,kw. | 1 |
| 38 | Western universali*.tw,kw. | 1 |
| 39 | Global pluriversali*.tw,kw. | 1 |
| 40 | Global south.tw,kw. | 869 |
| 41 | global north.tw,kw. | 403 |
| 42 | Intersectionality.tw,kw. | 1831 |
| 43 | Partnership ethic*.tw,kw. | 4 |
| 44 | Authorship ethic*.tw,kw. | 17 |
| 45 | exp *Racism/ or (*Prejudice/ or *Bias, Implicit/) | 17472 |
| 46 | Colonialism/ | 1605 |
| 47 | *Cultural Diversity/ | 7017 |
| 48 | *Social Justice/ | 6396 |
| 49 | racial equalit*.tw,kw. | 97 |
| 50 | (racial hierarch* or race hierarch*).tw,kw. | 75 |
| 51 | national hierarch*.tw,kw. | 9 |
| 52 | or/16-51 | 49681 |
| 53 | 15 and 52 | 1470 |
| 54 | limit 53 to (editorial or letter) | 117 |
| 55 | (view* or opinion* or thought* or discourse or idea* or think* or debate* or perspective* or definition* or terminology).tw,kw. | 1926203 |
| 56 | 53 and 55 | 438 |

**Embase 1974 to 2022 September 08**

| **#** | **Searches** | **Results** |
| --- | --- | --- |
| 1 | Global health.tw,kw. | 41030 |
| 2 | Global public health.tw,kw. | 9898 |
| 3 | global mental health.tw,kw. | 1241 |
| 4 | Transnational partnership*.tw,kw. | 7 |
| 5 | International partnership*.tw,kw. | 528 |
| 6 | World health.tw,kw. | 99021 |
| 7 | (Transnational cooperat* or transnational co-operat*).tw,kw. | 33 |
| 8 | transnational collaborat*.tw,kw. | 40 |
| 9 | International health.tw,kw. | 4540 |
| 10 | (International coopperat* or international co operat*).tw,kw. | 320 |
| 11 | international collaborat*.tw,kw. | 10810 |
| 12 | (Partnership* adj7 (LMIC* or HIC* or income countr*)).tw,kw. | 211 |
| 13 | *Global Health/ | 5342 |
| 14 | exp international cooperation/ | 218249 |
| 15 | or/1-14 | 333934 |
| 16 | Decoloni*.tw,kw. | 2502 |
| 17 | (Racism or racist).tw,kw. | 8046 |
| 18 | ((race* or racial*) adj3 (prejudic* or bias* or discriminat*)).tw,kw. | 4481 |
| 19 | (Oppress* adj5 (minorit* or people of colo?r or person* of colo?r or indigenous)).tw,kw. | 91 |
| 20 | (Oppress* adj5 (racial* or race*)).tw,kw. | 131 |
| 21 | (privilege* adj5 (racial* or race*)).tw,kw. | 103 |
| 22 | Whiteness.tw,kw. | 1079 |
| 23 | White privilege*.tw,kw. | 122 |
| 24 | white suprem*.tw,kw. | 160 |
| 25 | (white adj2 majorit*).tw,kw. | 1241 |
| 26 | colonial*.tw,kw. | 6867 |
| 27 | post-colonial*.tw,kw. | 253 |
| 28 | (Historical coloni* or historical coloni*).tw,kw. | 45 |
| 29 | ((high* income* countr* or HIC) adj3 domina*).tw,kw. | 18 |
| 30 | Northern ventriloquis*.tw,kw. | 1 |
| 31 | Epistem* violence.tw,kw. | 19 |
| 32 | Epistem* oppress*.tw,kw. | 4 |
| 33 | Epistem* injustice*.tw,kw. | 112 |
| 34 | epistem* knowledge*.tw,kw. | 16 |
| 35 | western epistem*.tw,kw. | 11 |
| 36 | indigenous epistem*.tw,kw. | 21 |
| 37 | Southern voice*.tw,kw. | 1 |
| 38 | Western universali*.tw,kw. | 0 |
| 39 | Global pluriversali*.tw,kw. | 0 |
| 40 | Global south.tw,kw. | 780 |
| 41 | global north.tw,kw. | 364 |
| 42 | Intersectionality.tw,kw. | 1809 |
| 43 | Partnership ethic*.tw,kw. | 4 |
| 44 | Authorship ethic*.tw,kw. | 18 |
| 45 | racial equalit*.tw,kw. | 80 |
| 46 | (racial hierarch* or race hierarch*).tw,kw. | 72 |
| 47 | national hierarch*.tw,kw. | 10 |
| 48 | exp *racism/ | 5208 |
| 49 | *cognitive bias/ or *implicit bias/ or *prejudice/ | 2311 |
| 50 | *colonialism/ | 202 |
| 51 | *cultural diversity/ | 1321 |
| 52 | *social justice/ | 4719 |
| 53 | or/16-52 | 35004 |
| 54 | 15 and 53 | 1242 |
| 55 | limit 54 to (editorial or letter) | 76 |
| 56 | (view* or opinion* or thought* or discourse or idea* or think* or debate* or perspective* or definition* or terminology).tw,kw. | 2435318 |
| 57 | 54 and 56 | 373 |

**Global Health**

| S19 | S17 AND S18 | 138 |
| --- | --- | --- |
| S18 | view* OR opinion* OR thought* OR discourse OR idea* OR think* OR debate* OR perspective* OR definition* OR terminology | 270,955 |

S17 limited to letters = 6 results

| S17 | S5 AND S16 | 406 |
| --- | --- | --- |
| S16 | S6 OR S7 OR S8 OR S9 OR S10 OR S11 OR S12 OR S13 OR S14 OR S15 | 6,954 |
| S15 | "racial equalit*" OR "racial hierarch*" or "race hierarch*" OR "national hierarch*" | 25 |
| S14 | "Global pluriversali*" OR "Global south" OR "global north" OR Intersectionality OR "Partnership ethic*" OR "Authorship ethic*" | 18 |
| S13 | "Northern ventriloquis*" OR "Epistem* violence" OR "Epistem* oppress*" OR "Epistem* injustice*" OR "epistem* knowledge*" OR "western epistem*" OR "indigenous epistem*" OR "Southern voice*" OR "Western universali*" | 907 |
| S12 | (("high* income* countr*" or HIC) N3 domina*) | 11 |
| S11 | colonial* OR "post-colonial*" OR ("Historical coloni*" or "historical coloni*") | 1,863 |
| S10 | Whiteness OR "White privilege*" OR "white suprem*" OR (white N2 majorit*) | 918 |
| S9 | (Oppress* N5 (racial* or race*)) OR (privilege* N5 (racial* or race*)) | 50 |
| S8 | (Oppress* N5 (minorit* or "people of colo#r" or "person* of colo#r" or indigenous)) | 17 |
| S7 | Racism or racist OR ((race* or racial*) N3 (prejudic* or bias* or discriminat*)) | 2,504 |
| S6 | Decoloni* | 975 |
| S5 | S1 OR S2 OR S3 OR S4 | 104,586 |
| S4 | DE "international cooperation" | 1,093 |
| S3 | (Partnership* N7 (LMIC* or HIC* or "income countr*")) | 68 |
| S2 | "Transnational cooperat*" or "transnational co-operat*" OR "transnational collaborat*" OR "International health" OR "International coopperat*" or "international co operat*" OR "international collaborat*" | 15,050 |
| S1 | "Global health" OR "Global public health" OR "global mental health" OR "Transnational partnership*" OR "International partnership*" OR "World health" | 90,943 |

**Scopus**

( TITLE-ABS-KEY ( decoloni* OR racism OR racist OR ( ( race* OR racial* ) W/3 ( prejudic* OR bias* OR discriminat* ) ) OR ( oppress* W/5 ( minorit* OR "people of color" OR "people of colour" OR "person* of color" OR "person of colour" OR indigenous ) ) OR ( oppress* W/5 ( racial* OR race* ) ) OR ( privilege* W/5 ( racial* OR race* ) ) OR whiteness OR "White privilege*" OR "white suprem*" OR ( white W/2 majorit* ) OR colonial* OR "post-colonial*" OR ( "Historical coloni*" OR "historical coloni*" ) OR ( ( "high* income* countr*" OR hic ) W/3 domina* ) OR "Northern ventriloquis*" OR "epistem* violence" OR "Epistem* oppress*" OR "Epistem* injustice*" OR "epistem* knowledge*" OR "western epistem*" OR "indigenous epistem*" OR "Southern voice*" OR "Western universali*" OR "Global pluriversali*" OR "Global south" OR "global north" OR intersectionality OR "Partnership ethic*" OR "Authorship ethic*" OR "racial equalit*" OR "racial hierarch*" OR "race hierarch*" OR "national hierarch*" ) ) AND ( TITLE-ABS-KEY ( "Global health" OR "Global public health" OR "global mental health" OR "Transnational partnership*" OR "International partnership*" OR "World health" OR "Transnational cooperat*" OR "transnational co-operat*" OR "transnational collaborat*" OR "International health" OR "International coopperat*" OR "international co operat*" OR "international collaborat*" ) OR TITLE-ABS-KEY ( ( partnership* W/7 ( lmic* OR hic* OR "income countr*" ) ) ) ) AND ( LIMIT-TO ( DOCTYPE , "ed" ) OR LIMIT-TO ( DOCTYPE , "le" ) ) = 85 results

( ( TITLE-ABS-KEY ( decoloni* OR racism OR racist OR ( ( race* OR racial* ) W/3 ( prejudic* OR bias* OR discriminat* ) ) OR ( oppress* W/5 ( minorit* OR "people of color" OR "people of colour" OR "person* of color" OR "person of colour" OR indigenous ) ) OR ( oppress* W/5 ( racial* OR race* ) ) OR ( privilege* W/5 ( racial* OR race* ) ) OR whiteness OR "White privilege*" OR "white suprem*" OR ( white W/2 majorit* ) OR colonial* OR "post-colonial*" OR ( "Historical coloni*" OR "historical coloni*" ) OR ( ( "high* income* countr*" OR hic ) W/3 domina* ) OR "Northern ventriloquis*" OR "epistem* violence" OR "Epistem* oppress*" OR "Epistem* injustice*" OR "epistem* knowledge*" OR "western epistem*" OR "indigenous epistem*" OR "Southern voice*" OR "Western universali*" OR "Global pluriversali*" OR "Global south" OR "global north" OR intersectionality OR "Partnership ethic*" OR "Authorship ethic*" OR "racial equalit*" OR "racial hierarch*" OR "race hierarch*" OR "national hierarch*" ) ) AND ( TITLE-ABS-KEY ( "Global health" OR "Global public health" OR "global mental health" OR "Transnational partnership*" OR "International partnership*" OR "World health" OR "Transnational cooperat*" OR "transnational co-operat*" OR "transnational collaborat*" OR "International health" OR "International coopperat*" OR "international co operat*" OR "international collaborat*" ) OR TITLE-ABS-KEY ( ( partnership* W/7 ( lmic* OR hic* OR "income countr*" ) ) ) ) ) AND ( TITLE-ABS-KEY ( view* OR opinion* OR thought* OR discourse OR idea* OR think* OR debate* OR perspective* OR definition* OR terminology ) ) = 429 results

**Google and Google Scholar**

decolonizing global health|views|opinions|thoughts|discourse|ideas|think|debate|perspectives|definitions|terminology

The search was undertaken on 08/09/22. The first 5 pages were screened and results (excluding journal articles because they duplicated those from the database search) were chosen.

**citationchaser(1)**

Pearl papers for backward and forward citation search:

1. Decolonising global health: if not now, when?

[https://gh.bmj.com/content/5/8/e003394?s=09](https://eur01.safelinks.protection.outlook.com/?url=https%3A%2F%2Fgh.bmj.com%2Fcontent%2F5%2F8%2Fe003394%3Fs%3D09&data=05%7C01%7CAnh.Tran%40ukhsa.gov.uk%7C56ae8081d08e426cac3e08daad2169cd%7Cee4e14994a354b2ead475f3cf9de8666%7C0%7C0%7C638012656279145240%7CUnknown%7CTWFpbGZsb3d8eyJWIjoiMC4wLjAwMDAiLCJQIjoiV2luMzIiLCJBTiI6Ik1haWwiLCJXVCI6Mn0%3D%7C3000%7C%7C%7C&sdata=n1%2F9fYpnoxb3kJFcfqoCnPbnb4pa55Wt3Z06flXw6qM%3D&reserved=0)

2. Decolonising global health in 2021: a roadmap to move from rhetoric to reform

[https://gh.bmj.com/content/6/3/e005604.abstract](https://eur01.safelinks.protection.outlook.com/?url=https%3A%2F%2Fgh.bmj.com%2Fcontent%2F6%2F3%2Fe005604.abstract&data=05%7C01%7CAnh.Tran%40ukhsa.gov.uk%7C56ae8081d08e426cac3e08daad2169cd%7Cee4e14994a354b2ead475f3cf9de8666%7C0%7C0%7C638012656279145240%7CUnknown%7CTWFpbGZsb3d8eyJWIjoiMC4wLjAwMDAiLCJQIjoiV2luMzIiLCJBTiI6Ik1haWwiLCJXVCI6Mn0%3D%7C3000%7C%7C%7C&sdata=SVbQf%2Bn97ZxBL41%2BfYkjAz%2FFpXwiExHIVW8%2B1C1exXI%3D&reserved=0)

3. Decolonising global health: transnational research partnerships under the spotlight

[https://academic.oup.com/inthealth/article-abstract/12/6/518/5962065](https://eur01.safelinks.protection.outlook.com/?url=https%3A%2F%2Facademic.oup.com%2Finthealth%2Farticle-abstract%2F12%2F6%2F518%2F5962065&data=05%7C01%7CAnh.Tran%40ukhsa.gov.uk%7C56ae8081d08e426cac3e08daad2169cd%7Cee4e14994a354b2ead475f3cf9de8666%7C0%7C0%7C638012656279145240%7CUnknown%7CTWFpbGZsb3d8eyJWIjoiMC4wLjAwMDAiLCJQIjoiV2luMzIiLCJBTiI6Ik1haWwiLCJXVCI6Mn0%3D%7C3000%7C%7C%7C&sdata=HnpDM0nqCU1iwEhbg0O2hMT1tE88PjPzTH%2FwWUNdoPQ%3D&reserved=0)

4. Decolonising global health: where are the Southern voices?

[https://gh.bmj.com/content/6/7/e006576.abstract](https://eur01.safelinks.protection.outlook.com/?url=https%3A%2F%2Fgh.bmj.com%2Fcontent%2F6%2F7%2Fe006576.abstract&data=05%7C01%7CAnh.Tran%40ukhsa.gov.uk%7C56ae8081d08e426cac3e08daad2169cd%7Cee4e14994a354b2ead475f3cf9de8666%7C0%7C0%7C638012656279145240%7CUnknown%7CTWFpbGZsb3d8eyJWIjoiMC4wLjAwMDAiLCJQIjoiV2luMzIiLCJBTiI6Ik1haWwiLCJXVCI6Mn0%3D%7C3000%7C%7C%7C&sdata=B%2FCZqTirr9ofs5WX4GLPIu1JpnultE%2BkUMoWsoL%2BxHQ%3D&reserved=0)

5. The words we choose matter: recognising the importance of language in decolonising global health

[https://www.thelancet.com/journals/langlo/article/PIIS2214-109X(21)00197-2/fulltext](https://eur01.safelinks.protection.outlook.com/?url=https%3A%2F%2Fwww.thelancet.com%2Fjournals%2Flanglo%2Farticle%2FPIIS2214-109X(21)00197-2%2Ffulltext&data=05%7C01%7CAnh.Tran%40ukhsa.gov.uk%7C56ae8081d08e426cac3e08daad2169cd%7Cee4e14994a354b2ead475f3cf9de8666%7C0%7C0%7C638012656279145240%7CUnknown%7CTWFpbGZsb3d8eyJWIjoiMC4wLjAwMDAiLCJQIjoiV2luMzIiLCJBTiI6Ik1haWwiLCJXVCI6Mn0%3D%7C3000%7C%7C%7C&sdata=403ZtTP2P1I38CizzeCpwHrJY260rv%2B2b%2ByOml9d4nU%3D&reserved=0)

6. Why and for whom are we decolonising global health?

[https://www.thelancet.com/journals/langlo/article/PIIS2214-109X(21)00317-X/fulltext](https://eur01.safelinks.protection.outlook.com/?url=https%3A%2F%2Fwww.thelancet.com%2Fjournals%2Flanglo%2Farticle%2FPIIS2214-109X(21)00317-X%2Ffulltext&data=05%7C01%7CAnh.Tran%40ukhsa.gov.uk%7C56ae8081d08e426cac3e08daad2169cd%7Cee4e14994a354b2ead475f3cf9de8666%7C0%7C0%7C638012656279145240%7CUnknown%7CTWFpbGZsb3d8eyJWIjoiMC4wLjAwMDAiLCJQIjoiV2luMzIiLCJBTiI6Ik1haWwiLCJXVCI6Mn0%3D%7C3000%7C%7C%7C&sdata=0j40Np%2BgqD1u1QmJJYiE22qKi%2BsW%2BoyRUgBpZfc1mww%3D&reserved=0)

7. The Myth of decolonising global health

[https://www.thelancet.com/journals/lancet/article/PIIS0140-6736(21)02428-4/fulltext](https://eur01.safelinks.protection.outlook.com/?url=https%3A%2F%2Fwww.thelancet.com%2Fjournals%2Flancet%2Farticle%2FPIIS0140-6736(21)02428-4%2Ffulltext&data=05%7C01%7CAnh.Tran%40ukhsa.gov.uk%7C56ae8081d08e426cac3e08daad2169cd%7Cee4e14994a354b2ead475f3cf9de8666%7C0%7C0%7C638012656279145240%7CUnknown%7CTWFpbGZsb3d8eyJWIjoiMC4wLjAwMDAiLCJQIjoiV2luMzIiLCJBTiI6Ik1haWwiLCJXVCI6Mn0%3D%7C3000%7C%7C%7C&sdata=jUH4BUConzVAdKsVbMBxUAHLqoL%2FuvPuJaPeCqKkGC0%3D&reserved=0)

Backward citation = 67 results forward citation = 191 results.

The citation search was undertaken on 24/10/22

1.	Haddaway NR, Grainger MJ, Gray CT. Citationchaser: an R package and shiny APP for forward and backward citations chasing in academic searching. Zenodo. 2021.

**Search two: August 2024**

**MEDLINE**

Ovid MEDLINE(R) Epub Ahead of Print and In-Process, In-Data-Review & Other Non-Indexed Citations and Daily <August 27, 2024>

| **#** | **Query** | **Results from 29 Aug 2024** |
| --- | --- | --- |
| 1 | Global health.tw,kw. | 46,119 |
| 2 | Global public health.tw,kw. | 12,377 |
| 3 | global mental health.tw,kw. | 1,471 |
| 4 | Transnational partnership*.tw,kw. | 9 |
| 5 | International partnership*.tw,kw. | 447 |
| 6 | World health.tw,kw. | 95,474 |
| 7 | (Transnational cooperat* or transnational co-operat*).tw,kw. | 32 |
| 8 | transnational collaborat*.tw,kw. | 40 |
| 9 | International health.tw,kw. | 4,350 |
| 10 | (International coopperat* or international co operat*).tw,kw. | 238 |
| 11 | international collaborat*.tw,kw. | 8,792 |
| 12 | (Partnership* adj7 (LMIC* or HIC* or income countr*)).tw,kw. | 222 |
| 13 | *Global Health/ | 23,433 |
| 14 | *Internationality/ or *International Cooperation/ or *International Educational Exchange/ | 28,778 |
| 15 | or/1-14 | 203,759 |
| 16 | Decoloni*.tw,kw. | 2,700 |
| 17 | (Racism or racist).tw,kw. | 10,893 |
| 18 | ((race* or racial*) adj3 (prejudic* or bias* or discriminat*)).tw,kw. | 5,196 |
| 19 | (Oppress* adj5 (minorit* or people of colo?r or person* of colo?r or indigenous)).tw,kw. | 127 |
| 20 | (Oppress* adj5 (racial* or race*)).tw,kw. | 185 |
| 21 | (privilege* adj5 (racial* or race*)).tw,kw. | 141 |
| 22 | Whiteness.tw,kw. | 1,631 |
| 23 | White privilege*.tw,kw. | 133 |
| 24 | white suprem*.tw,kw. | 267 |
| 25 | (white adj2 majorit*).tw,kw. | 767 |
| 26 | colonial*.tw,kw. | 8,377 |
| 27 | post-colonial*.tw,kw. | 278 |
| 28 | (Historical coloni* or historical coloni*).tw,kw. | 59 |
| 29 | ((high* income* countr* or HIC) adj3 domina*).tw,kw. | 23 |
| 30 | Northern ventriloquis*.tw,kw. | 1 |
| 31 | Epistem* violence.tw,kw. | 32 |
| 32 | Epistem* oppress*.tw,kw. | 7 |
| 33 | Epistem* injustice*.tw,kw. | 260 |
| 34 | epistem* knowledge*.tw,kw. | 22 |
| 35 | western epistem*.tw,kw. | 21 |
| 36 | indigenous epistem*.tw,kw. | 33 |
| 37 | Southern voice*.tw,kw. | 1 |
| 38 | Western universali*.tw,kw. | 2 |
| 39 | Global pluriversali*.tw,kw. | 1 |
| 40 | Global south.tw,kw. | 1,543 |
| 41 | global north.tw,kw. | 739 |
| 42 | Intersectionality.tw,kw. | 3,081 |
| 43 | Partnership ethic*.tw,kw. | 4 |
| 44 | Authorship ethic*.tw,kw. | 18 |
| 45 | exp *Racism/ or (*Prejudice/ or *Bias, Implicit/) | 19,201 |
| 46 | Colonialism/ | 1,747 |
| 47 | *Cultural Diversity/ | 7,312 |
| 48 | *Social Justice/ | 6,718 |
| 49 | racial equalit*.tw,kw. | 108 |
| 50 | (racial hierarch* or race hierarch*).tw,kw. | 102 |
| 51 | national hierarch*.tw,kw. | 12 |
| 52 | or/16-51 | 58,971 |
| 53 | 15 and 52 | 1,810 |
| 54 | limit 53 to (editorial or letter) | 142 |
| 55 | (view* or opinion* or thought* or discourse or idea* or think* or debate* or perspective* or definition* or terminology).tw,kw. | 2,192,731 |
| 56 | 53 and 55 | 554 |
| 57 | limit 56 to dt=20220901-20240829 | 115 |

**Embase**

Embase <1974 to 2024 August 28>

| **#** | **Query** | **Results from 29 Aug 2024** |
| --- | --- | --- |
| 1 | Global health.tw,kw. | 56,425 |
| 2 | Global public health.tw,kw. | 14,368 |
| 3 | global mental health.tw,kw. | 1,700 |
| 4 | Transnational partnership*.tw,kw. | 8 |
| 5 | International partnership*.tw,kw. | 612 |
| 6 | World health.tw,kw. | 118,831 |
| 7 | (Transnational cooperat* or transnational co-operat*).tw,kw. | 45 |
| 8 | transnational collaborat*.tw,kw. | 49 |
| 9 | International health.tw,kw. | 5,054 |
| 10 | (International coopperat* or international co operat*).tw,kw. | 329 |
| 11 | international collaborat*.tw,kw. | 12,873 |
| 12 | (Partnership* adj7 (LMIC* or HIC* or income countr*)).tw,kw. | 283 |
| 13 | *Global Health/ | 7,419 |
| 14 | exp international cooperation/ | 244,611 |
| 15 | or/1-14 | 394,613 |
| 16 | Decoloni*.tw,kw. | 3,368 |
| 17 | (Racism or racist).tw,kw. | 12,150 |
| 18 | ((race* or racial*) adj3 (prejudic* or bias* or discriminat*)).tw,kw. | 6,001 |
| 19 | (Oppress* adj5 (minorit* or people of colo?r or person* of colo?r or indigenous)).tw,kw. | 127 |
| 20 | (Oppress* adj5 (racial* or race*)).tw,kw. | 184 |
| 21 | (privilege* adj5 (racial* or race*)).tw,kw. | 151 |
| 22 | Whiteness.tw,kw. | 1,502 |
| 23 | White privilege*.tw,kw. | 143 |
| 24 | white suprem*.tw,kw. | 247 |
| 25 | (white adj2 majorit*).tw,kw. | 1,652 |
| 26 | colonial*.tw,kw. | 8,004 |
| 27 | post-colonial*.tw,kw. | 306 |
| 28 | (Historical coloni* or historical coloni*).tw,kw. | 50 |
| 29 | ((high* income* countr* or HIC) adj3 domina*).tw,kw. | 29 |
| 30 | Northern ventriloquis*.tw,kw. | 2 |
| 31 | Epistem* violence.tw,kw. | 32 |
| 32 | Epistem* oppress*.tw,kw. | 5 |
| 33 | Epistem* injustice*.tw,kw. | 244 |
| 34 | epistem* knowledge*.tw,kw. | 19 |
| 35 | western epistem*.tw,kw. | 19 |
| 36 | indigenous epistem*.tw,kw. | 32 |
| 37 | Southern voice*.tw,kw. | 1 |
| 38 | Western universali*.tw,kw. | 1 |
| 39 | Global pluriversali*.tw,kw. | 0 |
| 40 | Global south.tw,kw. | 1,386 |
| 41 | global north.tw,kw. | 702 |
| 42 | Intersectionality.tw,kw. | 3,090 |
| 43 | Partnership ethic*.tw,kw. | 4 |
| 44 | Authorship ethic*.tw,kw. | 20 |
| 45 | racial equalit*.tw,kw. | 98 |
| 46 | (racial hierarch* or race hierarch*).tw,kw. | 96 |
| 47 | national hierarch*.tw,kw. | 13 |
| 48 | exp *racism/ | 7,319 |
| 49 | *cognitive bias/ or *implicit bias/ or *prejudice/ | 2,852 |
| 50 | *colonialism/ | 305 |
| 51 | *cultural diversity/ | 1,609 |
| 52 | *social justice/ | 5,083 |
| 53 | or/16-52 | 45,881 |
| 54 | 15 and 53 | 1,683 |
| 55 | limit 54 to (editorial or letter) | 102 |
| 56 | (view* or opinion* or thought* or discourse or idea* or think* or debate* or perspective* or definition* or terminology).tw,kw. | 2,777,521 |
| 57 | 54 and 56 | 520 |
| 58 | limit 57 to dd=20220908-20240829 | 42 |

**Global Health**

Global Health <1973 to 2024 Week 34>

| **#** | **Query** | **Results from 29 Aug 2024** |
| --- | --- | --- |
| 1 | Global health.tw. | 16,271 |
| 2 | Global public health.tw. | 6,361 |
| 3 | global mental health.tw. | 184 |
| 4 | Transnational partnership*.tw. | 4 |
| 5 | International partnership*.tw. | 132 |
| 6 | World health.tw. | 45,102 |
| 7 | (Transnational cooperat* or transnational co-operat*).tw. | 10 |
| 8 | transnational collaborat*.tw. | 13 |
| 9 | International health.tw. | 1,968 |
| 10 | (International coopperat* or international co operat*).tw. | 61 |
| 11 | international collaborat*.tw. | 1,534 |
| 12 | (Partnership* adj7 (LMIC* or HIC* or income countr*)).tw. | 84 |
| 13 | exp international cooperation/ | 1,170 |
| 14 | or/1-13 | 69,656 |
| 15 | Decoloni*.tw. | 1,205 |
| 16 | (Racism or racist).tw. | 2,142 |
| 17 | ((race* or racial*) adj3 (prejudic* or bias* or discriminat*)).tw. | 2,739 |
| 18 | (Oppress* adj5 (minorit* or people of colo?r or person* of colo?r or indigenous)).tw. | 22 |
| 19 | (Oppress* adj5 (racial* or race*)).tw. | 40 |
| 20 | (privilege* adj5 (racial* or race*)).tw. | 25 |
| 21 | Whiteness.tw. | 863 |
| 22 | White privilege*.tw. | 18 |
| 23 | white suprem*.tw. | 41 |
| 24 | (white adj2 majorit*).tw. | 178 |
| 25 | colonial*.tw. | 2,085 |
| 26 | post-colonial*.tw. | 130 |
| 27 | (Historical coloni* or historical coloni*).tw. | 14 |
| 28 | ((high* income* countr* or HIC) adj3 domina*).tw. | 8 |
| 29 | Northern ventriloquis*.tw. | 0 |
| 30 | Epistem* violence.tw. | 3 |
| 31 | Epistem* oppress*.tw. | 0 |
| 32 | Epistem* injustice*.tw. | 20 |
| 33 | epistem* knowledge*.tw. | 1 |
| 34 | western epistem*.tw. | 7 |
| 35 | indigenous epistem*.tw. | 7 |
| 36 | Southern voice*.tw. | 0 |
| 37 | Western universali*.tw. | 2 |
| 38 | Global pluriversali*.tw. | 1 |
| 39 | Global south.tw. | 647 |
| 40 | global north.tw. | 268 |
| 41 | Intersectionality.tw. | 582 |
| 42 | Partnership ethic*.tw. | 2 |
| 43 | Authorship ethic*.tw. | 0 |
| 44 | racial equalit*.tw. | 11 |
| 45 | (racial hierarch* or race hierarch*).tw. | 16 |
| 46 | national hierarch*.tw. | 4 |
| 47 | or/15-46 | 8,876 |
| 48 | 14 and 47 | 364 |
| 49 | limit 48 to (editorial or correspondence) | 5 |
| 50 | (view* or opinion* or thought* or discourse or idea* or think* or debate* or perspective* or definition* or terminology).tw. | 295,311 |
| 51 | 48 and 50 | 143 |
| 52 | limit 51 to yr="2022 - 2024" | 68 |

**Scopus**

( TITLE-ABS-KEY ( decoloni* OR racism OR racist OR ( ( race* OR racial* ) W/3 ( prejudic* OR bias* OR discriminat* ) ) OR ( oppress* W/5 ( minorit* OR "people of color" OR "people of colour" OR "person* of color" OR "person of colour" OR indigenous ) ) OR ( oppress* W/5 ( racial* OR race* ) ) OR ( privilege* W/5 ( racial* OR race* ) ) OR whiteness OR "White privilege*" OR "white suprem*" OR ( white W/2 majorit* ) OR colonial* OR "post-colonial*" OR ( "Historical coloni*" OR "historical coloni*" ) OR ( ( "high* income* countr*" OR hic ) W/3 domina* ) OR "Northern ventriloquis*" OR "epistem* violence" OR "Epistem* oppress*" OR "Epistem* injustice*" OR "epistem* knowledge*" OR "western epistem*" OR "indigenous epistem*" OR "Southern voice*" OR "Western universali*" OR "Global pluriversali*" OR "Global south" OR "global north" OR intersectionality OR "Partnership ethic*" OR "Authorship ethic*" OR "racial equalit*" OR "racial hierarch*" OR "race hierarch*" OR "national hierarch*" ) ) AND ( TITLE-ABS-KEY ( "Global health" OR "Global public health" OR "global mental health" OR "Transnational partnership*" OR "International partnership*" OR "World health" OR "Transnational cooperat*" OR "transnational co-operat*" OR "transnational collaborat*" OR "International health" OR "International coopperat*" OR "international co operat*" OR "international collaborat*" ) OR TITLE-ABS-KEY ( ( partnership* W/7 ( lmic* OR hic* OR "income countr*" ) ) ) ) AND PUBYEAR > 2021 AND PUBYEAR < 2025 AND ( LIMIT-TO ( DOCTYPE,"ed" ) OR LIMIT-TO ( DOCTYPE,"le" ) ) = 65 documents

( ( TITLE-ABS-KEY ( decoloni* OR racism OR racist OR ( ( race* OR racial* ) W/3 ( prejudic* OR bias* OR discriminat* ) ) OR ( oppress* W/5 ( minorit* OR "people of color" OR "people of colour" OR "person* of color" OR "person of colour" OR indigenous ) ) OR ( oppress* W/5 ( racial* OR race* ) ) OR ( privilege* W/5 ( racial* OR race* ) ) OR whiteness OR "White privilege*" OR "white suprem*" OR ( white W/2 majorit* ) OR colonial* OR "post-colonial*" OR ( "Historical coloni*" OR "historical coloni*" ) OR ( ( "high* income* countr*" OR hic ) W/3 domina* ) OR "Northern ventriloquis*" OR "epistem* violence" OR "Epistem* oppress*" OR "Epistem* injustice*" OR "epistem* knowledge*" OR "western epistem*" OR "indigenous epistem*" OR "Southern voice*" OR "Western universali*" OR "Global pluriversali*" OR "Global south" OR "global north" OR intersectionality OR "Partnership ethic*" OR "Authorship ethic*" OR "racial equalit*" OR "racial hierarch*" OR "race hierarch*" OR "national hierarch*" ) ) AND ( TITLE-ABS-KEY ( "Global health" OR "Global public health" OR "global mental health" OR "Transnational partnership*" OR "International partnership*" OR "World health" OR "Transnational cooperat*" OR "transnational co-operat*" OR "transnational collaborat*" OR "International health" OR "International coopperat*" OR "international co operat*" OR "international collaborat*" ) OR TITLE-ABS-KEY ( ( partnership* W/7 ( lmic* OR hic* OR "income countr*" ) ) ) ) ) AND ( TITLE-ABS-KEY ( view* OR opinion* OR thought* OR discourse OR idea* OR think* OR debate* OR perspective* OR definition* OR terminology ) ) AND PUBYEAR > 2021 AND PUBYEAR < 2025 = 369 documents

**Google and Google Scholar**

decolonizing global health|views|opinions|thoughts|discourse|ideas|think|debate|perspectives|definitions|terminology

A custom date range was applied (2022-2024)

The search was undertaken on 28/08/2024. The first 5 pages were screened and results were chosen.
