## Supplementary material for "Decolonising Global Health: a scoping review": S2 Appendix - Author Locations Tables

**S2 Appendix: Location for authors of included papers**

**Table to accompany Fig 2: Location of authors contributing to papers included in the review.**

| **Author location*** | **Number of papers**** |
| --- | --- |
| USA | 53 |
| UK | 30 |
| Canada | 17 |
| Nigeria | 13 |
| Australia | 9 |
| Switzerland | 9 |
| Kenya | 8 |
| South Africa | 7 |
| Uganda | 7 |
| Ghana | 6 |
| India | 6 |
| Malaysia | 6 |
| Belgium | 5 |
| China | 5 |
| Germany | 5 |
| Sri Lanka | 5 |
| Tanzania | 5 |
| Brazil | 4 |
| Rwanda | 4 |
| Norway | 3 |
| Sweden | 3 |
| Chile | 2 |
| Malawi | 2 |
| Netherlands | 2 |
| Pakistan | 2 |
| Peru | 2 |
| Spain | 2 |
| Argentina | 1 |
| Bangladesh | 1 |
| Benin | 1 |
| Botswana | 1 |
| Cameroon | 1 |
| Costa Rica | 1 |
| Ethiopia | 1 |
| Ecuador | 1 |
| France | 1 |
| Gambia | 1 |
| Guatemala | 1 |
| Ireland | 1 |
| Italy | 1 |
| Laos | 1 |
| Lebanon | 1 |
| Mali | 1 |
| Mexico | 1 |
| Mozambique | 1 |
| Nepal | 1 |
| New Zealand | 1 |
| Palestine | 1 |
| Philippines | 1 |
| Puerto Rico | 1 |
| Samoa | 1 |
| Saudi Arabia | 1 |
| Senegal | 1 |
| Sierra Leone | 1 |
| UAE | 1 |
| Zimbabwe | 1 |

***** Note: definition of author location refers to the primary institution that the author is affiliated with at the time of publication.

**Note: author locations were counted per paper, rather than per author. Where a paper had more than one author from the same location, this location would be counted once.

**Table for Fig 3: Location of the first author of each paper with multiple authors included in the review.**

| **First author location***** | **Number of papers** |
| --- | --- |
| USA | 27 |
| UK | 8 |
| Nigeria | 5 |
| Australia | 4 |
| Malaysia | 4 |
| Belgium | 3 |
| Canada | 3 |
| Switzerland | 3 |
| Uganda | 3 |
| Kenya | 2 |
| Sweden | 2 |
| Unknown | 2 |
| Bangladesh | 1 |
| Chile | 1 |
| Germany | 1 |
| India | 1 |
| Laos | 1 |
| Nepal | 1 |
| Netherlands | 1 |
| Norway | 1 |
| Puerto Rico | 1 |

******* Note: this data only includes papers with more than one author
